# Systematic review and meta-aggregate analysis of mental health service users’ perspectives on romantic and intimate relationships and their support needs

**DOI:** 10.1101/2025.03.31.25324758

**Authors:** Aisling Smith O’Connor, Helen Killaspy, Sharon Eager, Eunice Wu, Brynmor Lloyd-Evans

**Affiliations:** Division of Psychiatry, University College London, United Kingdom

## Abstract

We conducted a systematic review and meta-aggregate synthesis to collate qualitative evidence about mental health service users’ views on their needs for romantic and intimate relationships and support measures. Four databases were searched in April 2024 (PROSPERO ID: CRD42024503997). Eligible studies involved (i) adult participants with any mental health condition who had received/were receiving care from mental health services; and (ii) had qualitative data specific to romantic/intimate relationship needs and support implications. Quality appraisal was conducted using the JBI Critical Appraisal Checklist for Qualitative Research. Data extraction and synthesis were conducted using meta-aggregation and the level of confidence in the synthesised findings was assessed using the GRADE-CERQual approach. We included 17 studies and extracted 100 specific findings. From these, 17 categories were created and grouped into four synthesised findings: (i) individuals with mental health conditions have a desire for romantic and intimate relationships; (ii) mental health conditions can directly hinder romantic and/or intimate relationships at the individual-level; (ii) psychosocial factors impact romantic and/or intimate relationships; and (iv) services are lacking and can act as obstacles for service users to engage in/discuss romance and/or intimacy. We found that service users had clear needs relating to romantic and intimate relationships, but experienced multiple challenges in meeting these needs. In general, mental health services tended to not provide support in this area. Implications for practice include: (i) recognising romantic and intimate relationship needs as part of routine mental health care; (ii) making sexual health and healthy relationship information available; (iii) offering spaces for group discussion and support; and (iv) facilitating social activities to meet others and build social skills and confidence.

## Introduction

Romantic and intimate relationships are integral to most people’s lives and are a determinant of health and wellbeing [1]. Romance and intimacy encompass romantic love, physical intimacy, and/or sexual activity, and these experiences can benefit people’s physical, social, and mental health [2]. Notably, the World Health Organisation (WHO) define health as “a state of complete physical, social and mental wellbeing”[3], with romantic and intimate relationships contributing to all three facets of “complete” health. Furthermore, the WHO state that “the sexual rights of all persons must be respected, protected and fulfilled” and emphasise that this requires “a positive and respectful approach to sexuality and sexual relationships”[3].

This stance is echoed by findings that romance and intimacy can have positive effects for the general population and specifically for those with mental health conditions [4,5].

Better mental health predicts entry into a romantic relationship [4], and having a good relationship is associated with improvements to mental health symptoms for individuals with mental illness [4]. In the general population, findings suggest that romantic connections promote self-expansion and lower depressive symptoms [6]. Relatedly, romantic loneliness has been associated with negative affect and suicidal ideation in the general population [7]. For people with serious mental illness, those with a partner had greater relationship satisfaction, which is related to reduced symptoms and higher mental wellbeing [8].

Moreover, service users themselves have considered emotional and/or sexual intimacy to be a key facilitator and indicator of recovery [9].

Ninety percent of patients with a psychotic disorder identified a need for sexual expression and 83% a need for intimate relationships [10] and, in a small survey, nearly all had been sexually active [11]. Furthermore, Warner et al. reported that 50% of patients had engaged in sexual activity on psychiatric wards [12]. There is therefore a strong case for mental health services to recognise people’s needs and wishes for romantic/intimate relationships, to support service users’ mental wellbeing and recovery.

Recent policy reform reflects this across health and care sectors. Building upon service-user movements and intellectual and learning disability directives [13], more attention to relationships and sexuality has been granted. In the UK, the Transforming Care Model [14] and the Supported Loving Toolkit by Choice Support [15] outline how to support individuals with learning disabilities to form and maintain romantic/intimate relationships. Moreover, personal relationships (including domestic and family relationships) have been highlighted as one of the eligible support needs that Local Authorities have a duty to assess and provide support for [16]. The Care Quality Commission [17] policy guidance on sexual safety through empowerment recommends that social care staff are open to talking to service users about their sexual needs and activity. This is to promote safety, sexual health, and support people in developing healthy, meaningful relationships.

Although promising, high-level policy support does not guarantee that such guidance affects practice. Recent initiatives to improve community mental health care have focused on employment, housing, living, and social support – as seen in the Community Mental Health Framework [18]. Romantic and intimate relationship support currently does not have a clear focus in mental health services, despite its documented benefits [4,5,8,9]. This may reflect a stigmatisation and ‘taboo’ surrounding intimacy and sexuality [19,20].

Mental health staff are often apprehensive about providing support with service users’ needs and wishes for romantic relationships. Many feel uncomfortable as they feel ill- equipped owing to a lack of training [21,22], or that discussing relationships may violate professional boundaries or be intrusive [23,24]. This underscores the importance of educating staff to develop their confidence, shift attitudes, and strategize how to best support with this need [22,24–26].

However, the risk of abuse and exploitation in this often vulnerable population cannot be overlooked [27,28]. Service users’ gender may contribute to practitioners’ concerns, with female service users sometimes seen as requiring more protection [29] and male service users needing more ‘containment’ [30]. Generally, mental health services tend to view sexual intimacy amongst service users through a lens of risk-aversion [31,32]. Other challenges for people with mental health conditions in developing fulfilling romantic/intimate relationships include: impaired social functioning [33], sexual dysfunction from symptoms and medication side effects [34], and lack of access to privacy [35–37].

Internalised and external stigma are also important [38,39]. A recent Australian survey of the general public found that mental illness was commonly thought of as a relationship “dealbreaker” suggesting social distance towards mental illness extends to include romantic relationships [40].

There remains a lack of resources to equip staff with the necessary knowledge and training to feel confident in talking to service users about romantic and intimate relationships. This includes assessing service users’ capacity to consent to romantic and intimate relationships [41,42], as well as embedding sexual health as part of routine mental health care [31]. This may be an important consideration for inpatient as well as community settings, as sexual activity on wards is not uncommon [43].

Support in finding a partner [22] and fulfilling intimacy needs typically remain unmet in practice [44]. This in part reflects a lack of established models and programmes of support. A 2024 review of interventions aimed at developing and maintaining relationships among people with mental illness emphasised a lack of interventions overall, with only one designed for single service users, and a shortage for people with serious mental illness [19]. Most publications on this topic are recent [45]. “Supported dating” has been suggested as one potential method of support [9], alongside practical interventions, including social activities and assistance using dating sites [9,45]. Practitioners have noted the need for education on safe dating behaviours [23], given the risks of exploitation. These recommendations align with the finding that mental health service users were willing and capable of participating in safe sex interventions [46].

In the last five years, there has been a greater focus on positioning those with mental health conditions at the centre of this topic. To our knowledge, four previous reviews have been published on service users’ perspectives about sexuality and romantic/intimate relationships. These have included: a quantitative review focused on individuals with psychotic disorders [37]; an integrative review focused on individuals being treated under a forensic order [25]; and two qualitative reviews focused on individuals with serious mental illness [48,49]. Our review aimed to build upon McCann’s 2019 qualitative systematic review [48] and Hortal-Mas et al.’s 2022 qualitative meta-synthesis [49] to include studies relating to all mental health conditions. We chose to focus more specifically on service users’ needs for and experience of support from mental health services with romantic/intimate relationships, rather than their experience and understanding of their sexuality. Our review aimed to provide an up-to-date synthesis of service users’ reported needs for and views about support with romantic/intimate relationships. We focused on direct reports of service users’ views about support from mental health services, not authors’ inferences about what might be helpful. Through collating the available qualitative data, the primary aim was to inform healthcare practice and development on supporting mental health service user’s needs for romantic and intimate relationships, from a user-centred perspective.

## Methods

### Protocol and registration

This study’s protocol was pre-registered to PROSPERO on 14/03/2024 (CRD42024503997). We followed the 2020 PRISMA Checklist (S1 Table) and adhered to the PRISMA reporting guidelines [50].

### Eligibility criteria

#### Participants

Inclusion criteria were studies involving individuals aged 18 and over with a mental health condition who had received/were receiving care from mental health services.

#### Phenomena of interest

We aimed to explore the views of service users on their needs for romantic and/or intimate relationships, and how they can be best supported with this need by mental health services. Eligible papers related to relationships defined by sexual activity, romantic love, or increased physical intimacy, encompassing monogamy and non-monogamy.

#### Context

Eligible studies were those with participants who had current or past experience of mental health services, whether provided by statutory health and social care services or in the independent/private or voluntary sector.

#### Types of studies

Eligible studies were qualitative interviews, focus groups, papers reporting open-ended questionnaires, or surveys with free text responses. Ethnographic studies on mental health practice and single case studies were not included, to maintain our focus on service users’ direct reports of their experiences. There were no restrictions on the country or setting of the study or date of publication. The search was limited to peer-reviewed publications written in English language.

#### Search strategy

We searched terms relating to four concepts: romantic and intimate relationships; mental health; qualitative research; and service users. Synonyms for these core elements were generated and combined using Boolean operators. The searches were set to identify studies that included all key concepts in the title or abstract of studies. Necessary adjustments were made to adapt to the specifications of each database (S2 Table). The search was conducted on 09/04/2024. Of the included papers, forward and backward citation searches were undertaken using citation chaser software [51].

#### Data sources

The following electronic databases were searched: MEDLINE, PsycINFO, CINAHL, and Web of Science. A further search was explored using Google Scholar, where we searched “mental health” and “romantic/intimate relationships”. We chose to examine a minimum of the first 10 pages of results and to continue reviewing retrieved papers until they were no longer relevant to our research question.

#### Screening

An electronic systematic reviewing software, Covidence [52], was used to import, manage, compile, and evaluate the gathered studies. The search results were deduplicated by Covidence’s software prior to commencing screening. All titles and abstracts were screened by one reviewer (ASOC) and 10% of randomly selected titles and abstracts were independently screened by a second reviewer (EW). Once title and abstract screening was completed, all papers included for full text review were read by two independent raters (ASOC and EW). Reasons for exclusion were noted, under the categories of wrong patient population, wrong study design, or insufficient qualitative data (e.g. where romantic/intimate experiences were discussed but did not explore relationship needs or support implications). Conflicts were resolved through discussion among the two researchers involved in screening.

#### Data extraction

Data extraction was performed by one reviewer (ASOC). A data extraction table was created to extract relevant information from the selected studies. Extracted information included author, publication year, location, setting, phenomena of interest, methodology and analysis, lived-experience involvement, sample size, age range, gender, and participant diagnoses. The Joanna Briggs Institute (JBI) core definitions in meta-aggregative reviews [53] was followed to compile ‘findings’ (a verbatim extract of the authors interpretation of their results) and their accompanying ‘illustrations’ (a direct quotation of supporting data from the paper) from all studies. Each finding was assigned a level of credibility: unequivocal (beyond reasonable doubt and therefore not open to challenge); credible (lacking clear association with it and therefore open to challenge); or unsupported (findings not supported by data).

#### Methodological quality assessment

We used the JBI Critical Appraisal Checklist for Qualitative Research to assess the methodological quality of included studies [54]. This checklist involves 10 questions covering several domains, including ethical considerations, suitability of methodology, congruity of methodology and data collection, suitability of analysis, sources of biases, and validity of findings/interpretations.

#### Data analysis and synthesis

We followed the JBI Manual for Evidence Synthesis detailing a meta-aggregate approach [53]. Meta-aggregation was selected as it is sensitive to representing primary author’s findings without seeking to re-interpret them [55]. This involved pooling two or more similar findings to generate a category. Findings assessed as unsupported were not used to develop categories. Categories were created by developing an explanatory statement that conveyed the whole, inclusive meaning of findings. This grouping was similar to procedures used in basic qualitative methods, such as thematic analysis [56]. These categories were further aggregated to produce a synthesised finding that encapsulated the overarching meaning of a group of at least two similar categories. Synthesised findings were expressed as indicatory statements, with the view of generating descriptions that could be used to generate policy recommendations and service development [57].

#### Assessment of confidence in synthesised findings

We used the GRADE-Confidence in the Evidence from Reviews of Qualitative research approach (GRADE-CERQual; 55) to assess the level of confidence in the synthesised findings. This assessment is based on methodological limitations, coherence (the fit between the data from the primary studies and the review finding), adequacy (richness of data supporting a review finding), and relevance (applicability of data from the primary studies to the context specified in the review question). Following assessing each component, we rated the overall confidence in the culminated evidence supporting the synthesised finding as high, moderate, low, or very low.

#### Positionality statement

We selected meta-aggregation to minimise undue subjectivity in developing the findings. The meta-aggregation was performed independently by the lead author (ASOC), a heterosexual, cisgender woman who is in a relationship. This individual nature of the aggregation may have influenced how findings were interpreted. Reflexivity was supported by discussing the identified categories and synthesised findings with co-authors.

## Results

### Search results

A PRISMA flow diagram was created depicting the screening and extraction process, according to the predefined inclusion criteria (Fig 1). Of the 10% of studies that were double screened, there was a lack of consensus on inclusion/exclusion in 3% (15/503) that were resolved through discussion. There were 71 studies included for full text review, all of which were double screened. From these, 17 studies were included. Forward and backward citation searching of these 17 articles identified 694 additional papers (132 duplicates) of which 36 were double screened as full-text papers but none were included. Overall, inter- rater reliability was moderate for full text screening (Cohen’s kappa = 0.68), with nine conflicts out of 107 decisions and 91% agreement between raters.

**Fig 1.**
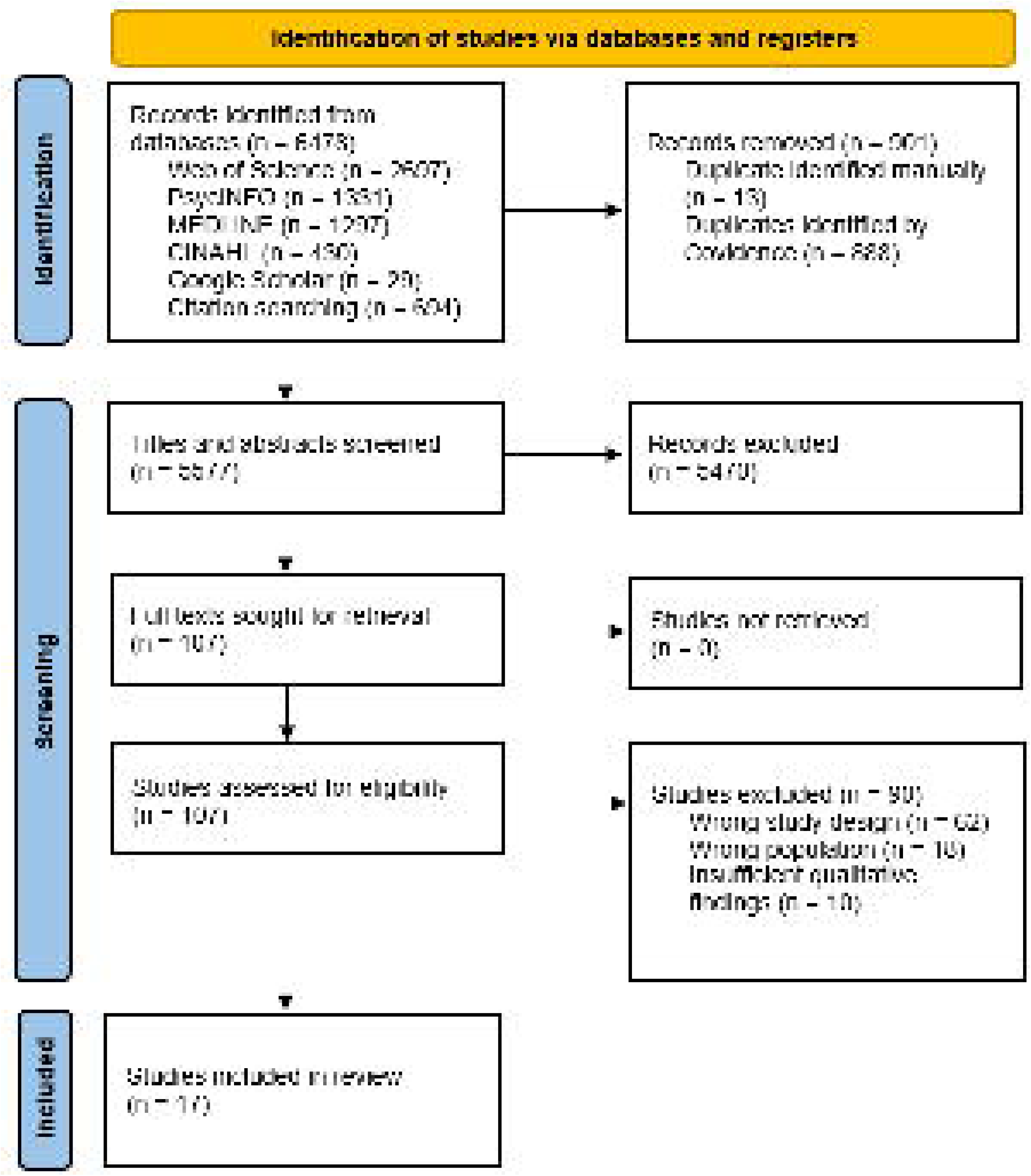
PRISMA flow diagram of study selection.

### Methodological Quality

We used the JBI Critical Appraisal Checklist for Qualitative Research (S3 Table) to appraise methodological quality of the 17 included studies. Two of the 10 questions had a lower number of “yes” responses: three papers (18%) had a statement locating the researcher culturally or theoretically (question six), and roughly half (53%) addressed the influence of the researcher on the research (question seven).

### Data extraction

Key study characteristics of included studies are summarised in Table 1. All papers were published after 2010, with seven of the 17 published since 2020. The majority of studies were conducted in high-income countries, with one from China and one from Iran.

**Table 1.** Characteristics of included studies.

| Author, publication year, and location | Setting | Phenomena of interest | Methodology and analysis | Lived-experience involvement | Participant demographics and diagnoses | Critical appraisal rating |
| --- | --- | --- | --- | --- | --- | --- |
| Brand et al., 2022 [33]<br>Australia | Forensic mental health unit | The sexuality and sexual experiences of forensic mental health participants treated under the requirements of a forensic order | Semi-structured interview;<br>Thematic analysis | None reported | N=14<br>Aged 25-61 (M=45.14)<br>11 males, 3 females<br>Schizophrenia, bipolar, unspecified psychosis | 8/10<br>High |
| Brown et al., 2014 [30]<br>United Kingdom | Forensic mental health unit | To examine how the public exclusion of sexuality from forensic unit results in sexuality being experienced as sectioned off or amputated | Semi-structured interview;<br>Thematic analysis | None reported | N=20<br>15 males, 5 females<br>A psychotic disorder, severe depression | 9/10<br>High |
| Budziszewska et al., 2020 [59]<br>Poland | Inpatient and outpatient psychiatric clinic | To examine how the illness has influenced their emotional experiences regarding love and their intimate relationship experiences, with an explicit intention to give voice to the patient's perspective | Semi-structured interview;<br>Interpretative phenomenological analysis | None reported | N=10<br>Aged 28-40<br>5 males, 5 females<br>Schizophrenia (6 outpatients, 4 inpatients) | 10/10<br>High |
| deJager et al., 2017 [60]<br>Netherlands | Community mental health care | To explore which problems participants encounter in establishing intimacy and maintaining intimate relationships | Semi-structured interview;<br>Grounded theory | None reported | N=28<br>Aged 22-62 (M=42)<br>19 males, 9 females<br>Psychotic disorder receiving flexible assertive community treatment | 7/10<br>High |
| Forrester-Jones et al., 2023 [61]<br>United Kingdom | Mental health charities and a mental health self-help network | To explore how people with mental health problems experience romantic relationships and what support they might need | Focus groups;<br>Thematic analysis | Approach was informed by discussions with an advisory group made up of one person with mental health problems | N=10<br>8 males, 2 females<br>Do not specify mental health diagnoses | 9/10<br>High |
| Kaplan et al., 2022 [62]<br>Israel | Psychiatric unit | To examine the sexual needs and challenges of people with severe mental illnesses admitted to an isolated psychiatric ward | Semi-structured interview;<br>Thematic analysis | None reported | N=13<br>Aged 32-67 (M=46.03)<br>13 males<br>Schizophrenia, psychotic disorders, or bipolar disorder | 9/10<br>High |
| Landi et al., 2020 [63]<br>Italy | Psychiatric residential setting | To better understand how sexuality and affectivity are expressed by service users within a psychiatric residential settings | Focus groups;<br>Content analysis | None reported | N=13<br>Aged >18<br>Psychotic disorders, personality disorders, bipolar disorder, depression | 9/10<br>High |
| McCann, 2010 [13]<br>United Kingdom | A community-based clinic in London that provided antipsychotic depot injections | To capture in-depth perspectives of people with a medical diagnosis of schizophrenia regarding intimate relationships | Semi-structured interview;<br>Content analysis data reduction | Development of interview topic guide included service user input | N=30<br>Aged 22-57 (M=41)<br>15 males, 15 7/females<br>Schizophrenia, schizotypal and delusional disorder | 7/10<br>High |
| Mizock et al., 2019 [64]<br>California | A psychosocial recovery centre | To better understand women with serious mental illness's experiences with romantic and intimate relationships | Semi-structured interview;<br>Grounded theory | None reported | N=20<br>Aged 32-66 (M=50)<br>20 females<br>Major depressive disorder, PTSD, anxiety, bipolar, borderline personality disorder, schizophrenia, | 9/10<br>High |

|  |  |  |  |  | schizoaffective |  |
| --- | --- | --- | --- | --- | --- | --- |
| Ostman, 2014 [65]<br>Sweden | Outpatient psychiatric services | To determine how people with serious mental illness experience sex and assess satisfaction with it in a broader evaluation of quality of life | In-depth interview;<br>Thematic analysis | Proposed interview questions were validated in collaboration with a service user research advisory group | N=20<br>Aged 33-82 (M=50)<br>16 males, 4 females<br>Do not specify diagnoses: all were currently in treatment in psychiatric out-patient services with serious mental illness | 10/10<br>High |
| Quinn & Happell, 2016 [66]<br>Australia | Secure forensic hospital | To explore perceptions of nurses and patients regarding sexual intimacy in a long-term mental health unit | Semi-structured interview;<br>Thematic analysis | None reported | N=10<br>Aged 25-48 M=35)<br>6 males, 4 females<br>Do not specify diagnoses: forensic inpatients on custodial supervision orders | 8/10<br>High |
| Quinn & Happell, 2015 [67]<br>Australia | Secure forensic hospital | To elicit the views of consumers and nurses about forming sexual relationships within this long-term and secure setting | In-depth interview;<br>Thematic analysis | None reported | N=10<br>Aged 25-48 (M=35)<br>6 males, 4 females<br>Do not specify diagnoses: forensic inpatients on custodial supervision orders | 8/10<br>High |
| Quinn & Happell, 2015 [36]<br>Australia | Secure forensic hospital | To explore perceptions of privacy and dignity for sexual relationships in a forensic mental health hospital | In-depth interview;<br>Thematic analysis | None reported | N=10<br>Aged 25-48 (M=35)<br>6 males, 4 females<br>Do not specify diagnoses: forensic inpatients on custodial supervision orders | 8/10<br>High |
| Raisi et al., 2018 [68]<br>Iran | Inpatient psychiatric hospital | To explore views of patients with severe mental illness and their care providers about sharing sexual problems with care providers in these patients within the context of Iran | Semi-structured interview;<br>Content analysis data reduction | None reported | N=4<br>Aged 17-49 (M=33)<br>Do not specify diagnoses further than having serious mental illnesses | 9/10<br>High |
| Robertson et al., 2015 [69]<br>United Kingdom | Inpatient psychiatric hospital | To explore the psychiatric inpatient experiences of lesbian and gay service users in relation to their intimate relationship needs and how these experiences affected their mental health recovery | Semi-structured interview;<br>Interpretative phenomenological analysis | Interview topic guide was developed in collaboration with a gay service user | N=6<br>Aged 31-57 (M=45)<br>3 males, 3 females<br>Schizophrenia, bipolar affective disorder and personality disorders | 9/10<br>High |
| White et al., 2021 [70]<br>United Kingdom | Community-based mental healthcare services | To investigate how people with experience of psychosis conceptualise romantic relationships and what support they would like in this area of their lives | Semi-structured interview;<br>Thematic analysis | None reported | N=10<br>Aged 21-64<br>6 males, 4 females<br>Experience of psychosis | 8/10<br>High |
| Yang et al., 2023 [71]<br>China | Inpatient and outpatient psychiatric hospital | To explore the sexual needs of people with schizophrenia and identify factors hindering sexual activities | Semi-structured interview;<br>Descriptive phenomenological approach | None reported | N=20<br>Aged 23-52 (M=36.9)<br>10 males, 10 females<br>Schizophrenia | 8/10<br>High |

N = sample size; M = mean.

Four studies included people with lived-experience of mental health problems in the research team. The studies were all relatively small, with sample sizes that ranged from four to 30 participants. Most studies focused on people with serious mental illness (n = 13), with three collecting data from forensic inpatients without specifying their mental health conditions. Nine studies involved inpatient populations, six were conducted with people living in the community, and two involved a mixture of inpatient and outpatient participants. Two studies involved focus groups and the remaining 15 used individual interviews, of which 12 were described as semi-structured and three as in-depth interviews. Analytic techniques included thematic analysis (n = 9), content analysis (n = 3), interpretative phenomenological analysis (n =2), grounded theory (n =2), and descriptive phenomenological analysis (n = 1). A total of 100 findings and accompanying illustrations were extracted from the included papers (see S4 Table).

### Review findings

#### Overview

The 100 extracted findings were aggregated into 17 categories (see S5 Table for procedural details) that were in-turn grouped into four synthesised findings (Table 2). The first synthesised finding (‘individuals with mental health conditions have a desire for romantic and intimate relationships’) had three categories composed of 14 findings of which 93% were rated as having ‘unequivocal’ levels of credibility. The second synthesised finding (‘mental health conditions directly hinder romantic and/or intimate relationships at the individual-level’) was drawn from three categories comprised of 18 findings, all of which were rated as ‘unequivocal’. The third synthesised finding (‘psychosocial factors impact romantic and/or intimate relationships’) was aggregated from five categories comprising 24 extracted findings, with 67% of these rated as ‘unequivocal’. The fourth synthesised finding (‘services are lacking and act as obstacles for service users to engage in romance and/or intimacy and its discussion’) was derived from six categories amalgamated from 44 findings, of which 75% were rated ‘unequivocal’.

**Table 2.**
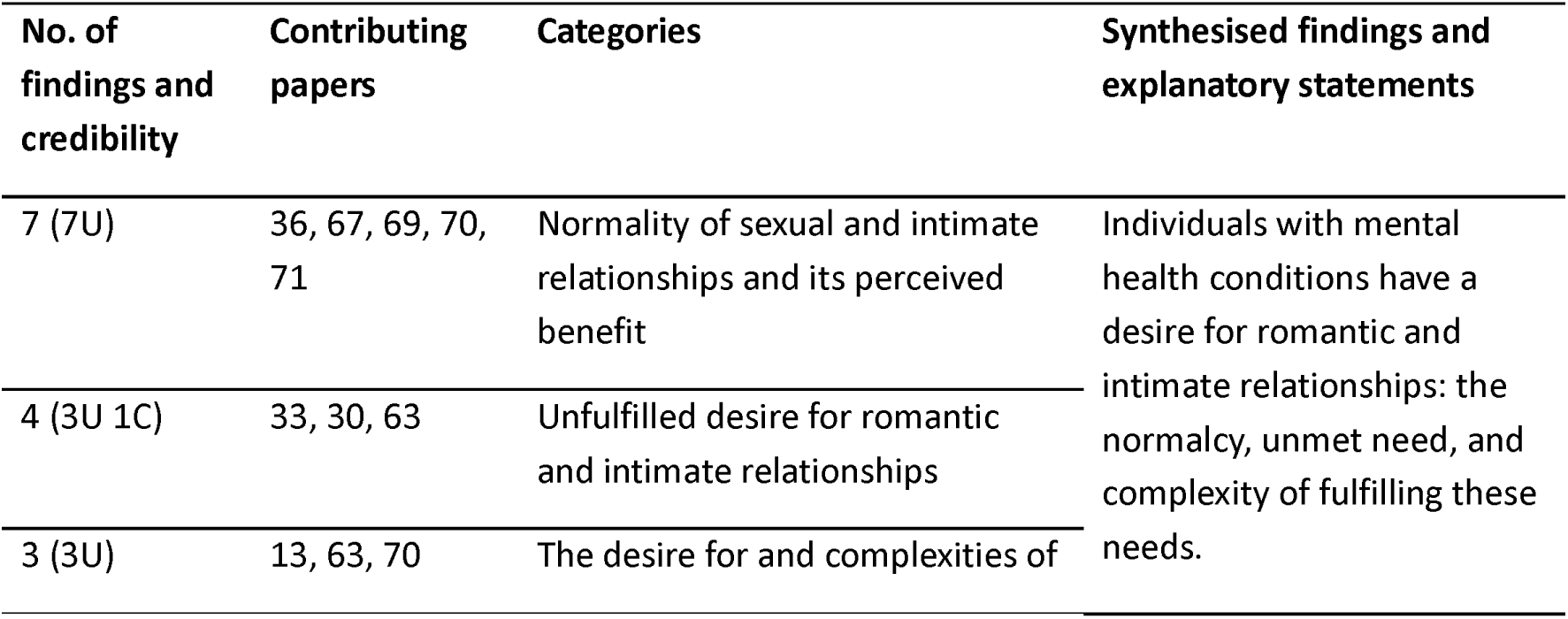
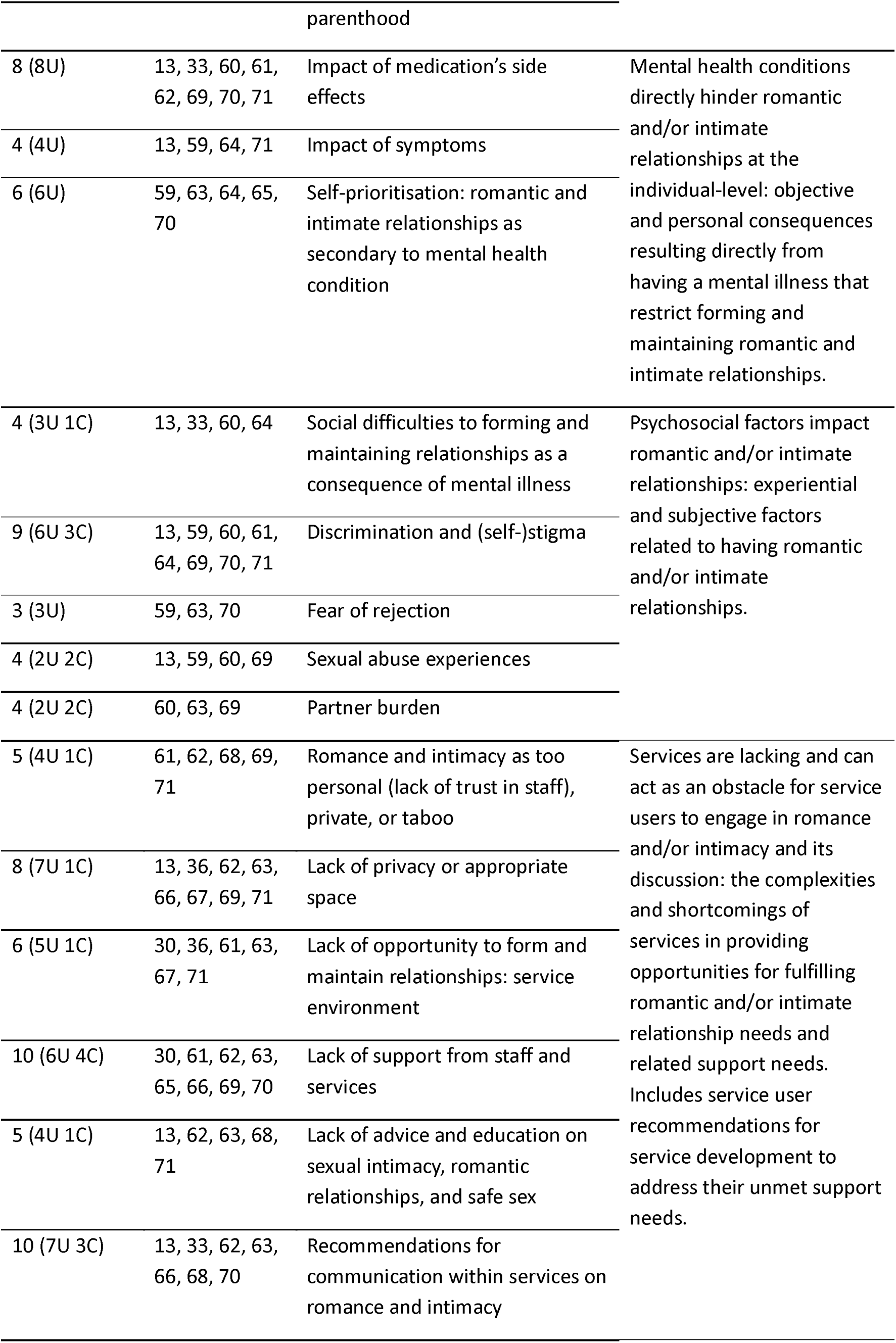
Summary of categorisation and synthesis formation.

We used GRADE-CERQual to establish the level of confidence to place in each synthesised finding (Table 3). No serious concerns were identified for any CERQual criterion. Findings were rated as having moderate confidence overall where there were minor concerns for most of the CERQual categories (rather than ‘no’ or ‘very minor’ concerns), or moderate concerns for any category. See S6 File for further details.

**Table 3.** GRADE-CERQual ratings for each synthesised findings.

| Synthesised finding | Papers contributing | GRADE-CERQual Assessment of Confidence in Findings |  |  |  |  |
| --- | --- | --- | --- | --- | --- | --- |
|  |  | Methodological limitations | Coherence | Adequacy | Relevance | Overall confidence |
| Individuals with mental health conditions have a desire for romantic and intimate relationships | 13, 30, 33, 36, 63, 67, 69, 70, 71 | No or very minor concerns | Minor concerns | No or very minor concerns | Minor concerns | High |
| Mental health conditions directly hinder romantic and/or intimate relationships at the individual-level | 13, 33, 59, 60, 61, 62, 63, 64, 65, 69, 70, 71 | No or very minor concerns | No or very minor concerns | No or very minor concerns | Minor concerns | High |
| Psychosocial factors impact romantic and/or intimate relationships | 13, 33, 59, 60, 61, 63, 64, 69, 70, 71 | No or very minor concerns | Minor concerns | Minor concerns | Minor concerns | Moderate |
| Services are lacking and act as obstacles for service users to engage in romance and/or intimacy and its discussion | 13, 30, 33, 36, 61, 62, 63, 65, 66, 67, 68, 69, 70, 71 | No or very minor concerns | No or very minor concerns | No or very minor concerns | Moderate concerns | Moderate |

### Individuals with mental health conditions have a desire for romantic and intimate relationships

#### Normality of sexual and intimate relationships and its perceived benefit

Studies emphasised the importance service users placed on romantic and intimate relationships. Having such relationships was seen as a normal human behaviour [36,67] that in-turn allowed service users feel socially included [69].

> “I think romantic relationships are incredibly important for anyone regardless of whether you have a mental health problem or not” [70].

> “It just felt like I had some normal.”[67].

Romantic relationships along with positive sexual experiences were considered an effective means of emotional regulation [67,71]. Moreover, being in a relationship was seen as aiding their wellbeing, providing “a sense of self-worth, a sense of identity, [and] a sense of self-purpose” [70]. Similarly, the “sense of feeling loved by another person” was identified as instrumental to progress and recovery [67].

### Unfulfilled desire for romantic and intimate relationships

Three studies highlighted service users’ unfulfilled desire for romance and intimacy [26,30,63].

> “I associate it with just general vibrancy… And life force and a dynamic that’s gone missing it’s disappeared” [30].

This absence resulted in feelings of loss and loneliness for participants [30,60]. This sentiment was followed with a hope for “a woman, a girlfriend” [63] or “to get married one day” [26].

### The desire and complexities of becoming a parent

In three studies, participants discussed the link between romantic and intimate relationships and parenthood [13,63,70]. These findings featured participants’ desire for having children and placing importance on having a family, while acknowledging the potential risks involved.

> “You know, love, connection and many other things, having children. I mean I dunno if I’ll be able, I hope to be able to have children… it’s very important to me”[70].

These risks involved “a nightmare situation” of service user’s children being placed in the care of social services [13,70] or a fear of potentially harming their child [70]. These risks outweighed the desire for having children, which inhibited some from becoming a parent.

> “At the moment because of my illness – in case I get sick and they get put into a home… no children” [13].

One participant [63] maintained a desire to have children, but their parents and mental health professionals advised “that it was better to take the [contraceptive] pill.”

### Mental health conditions directly hinder romantic and/or intimate relationships at the individual-level

#### Impact of medication side effects

Studies consistently identified medication side effects as a barrier to engaging in physical intimacy. Erectile dysfunction [33,60,62,71], vaginal dryness [13], difficulty in orgasming [69], and a lack of interest in sex were noted as deterrents [61].

> “When I have sex with my lover, sometimes I cannot get an erection. Even if I have an erection, it does not last for a while” [71].

Notably, one participant [13] contradicted this finding, attesting that their “sex drive went super high” when prescribed fluoxetine/Prozac. Additional side effects such as weight gain and sweating were also acknowledged [70]. This negatively affected their confidence, however this was a peripheral barrier rather than a direct hindrance, and was referenced less.

#### Impact of symptoms

Four papers identified participants’ mental health symptoms as an enduring factor detrimental to romantic or intimate involvement [13,59,64,71]. Changes in the experience of love due to symptom interference was cited as a barrier [59], as was an inability to orgasm due to emotional withdrawal [13] and difficulty concentrating [64].

An apparent awareness of the vulnerability that accompanies psychotic symptoms was also acknowledged, leading to sexual inactivity:

> “I sometimes have an erection in the morning but I do not care. Because at the moment I am out of control I’m not too… I’m not very trusting in myself to do anything else” [71].

#### Self-prioritisation

The absence of a desire for romantic and intimate relationships was a recurrent finding. Mental illness was deemed as all-consuming for many, resulting in not feeling the need for intimacy [63] or ready for a relationship [59]. Voluntary abstinence was common as a means of prioritising symptom management [59,64,65,71].

> “What dating life? I’m too busy: trying to be - it’s all I can do to take care of myself” [64].

### Psychosocial factors impact romantic and/or intimate relationships

#### Social difficulties to forming and maintaining relationships as a consequence of mental illness

Social difficulties manifested as being socially withdrawn [33] and lacking social skills [13,60], negatively impacting relationship initiation. Additionally, service users struggled to find a prospective partner with a similar level of functioning or someone socially compatible [13,64]:

> “I look at girls in the street but don’t know what to do. The kind of girls I meet in the drop-in… they are having a hard time in their heads. That’s not the kind of girl I want”[13].

However, one participant countered this:

> “I am seeing a woman in the hostel. We have been together for three years. She has schizophrenia and learning difficulties. We want to get married and live together”[13].

#### Discrimination and stigma

A recurring barrier to forming and sustaining relationships was discrimination and stigma. Social stigma and low status were viewed as reasons for being perceived as unfit for relationships [13,59–61,64,70,71].

> “Unfortunately he found out I’m in [name of hospital] and he didn’t want to be with me any more” [59].

> “I was thinking: you will lose him for sure. Who would want a psychosis?” [60]

The findings were also intertwined with self-stigma [13,60,61,64]. Participants referred to themselves as “a psychosis” [60] or “a bipolar” [64]. One participant exhibited a preference for “a normal person” as they “wouldn’t like to have a relationship with another schizophrenic”, demonstrating the identity-first language and internalised stigma [61].

#### Fear of rejection

Three studies identified fear of rejection as a barrier to initiating a relationship [59,63,70]. This fear was associated with worsening one’s emotional state:

> “If you are also lovesick this is really a bad suffering” [63].

#### Sexual abuse experiences

Four studies identified participants’ experiences of sexual abuse [13,59,60,69]. As a result, pursuing sexual relationships was negatively affected, with some requiring “a relationship with no sex life” [59]. One participant [69] wished to access talking therapy a to process previous abusive experiences:

> “I don’t talk to my workers about anything… for what I’ve been through in my time it’s not an injection I need, its counselling, it’s more support, more talkin’.”

#### Partner burden

The feeling of burdening partners was also discussed [60,63,69].

There was a sense of culpability due to pressures and expectations of partners to be supportive [60,69]. Additionally, guilt over unfavourable actions toward partners was noted as “the biggest barrier to recovery ” [69].

Partners’ relationship management difficulties were also highlighted. Some participants noted that their partners became overwhelmed and ended relationships [63]. Furthermore, partners’ mental wellbeing was also negatively influenced, compounding feelings of guilt:

> “He just really wanted to help me… At a certain point he became very depressed. I felt and still feel extremely guilty about it” [60].

### Services are lacking and may act as obstacles for service users to engage in and discuss romance and/or intimacy

#### Romance and intimacy as too personal (lack of trust in staff), private, or taboo

Several factors discouraged discussing romance and intimacy within a mental health service. Personal attitudes towards discourse about relationships, such as shame [69] and the perception that it is “still too private” [71], resulted in an unwillingness to talk about this with staff. Furthermore, service users acknowledged contextual barriers within a mental health setting, such as concerns that discussing the topic would cross limits or be ‘taboo’ [62,68].

> “Really maybe if there was a little bit more reference to the topic… it would light things up more positively rather than being such a taboo”[62].

Participants noted that discussing romance may not be within staff’s remit, accepting that some staff members may not consider it appropriate [68] and that romance may be a private topic to mental health staff themselves [61]. Moreover, some participants believed that the power imbalance between professionals and themselves was incompatible with discussing romantic/intimate relationships, where trust is required [70].

#### Lack of privacy or an appropriate space

In eight studies, participants discussed the absence of appropriate, private spaces as a problem in inpatient settings, but not one which always precluded sexual activity [36,62,63,66,67,71]. Service users described having to “go to extremes of finding some secret spot” [36], often resulting in feelings of discomfort [36,63,71] and a lack of dignity [66,67]. Being under the care of staff was marked by constant supervision and unpredictable ‘alone time’ [13,62], leading to “never [having] any real sense of being able to have… the relationship” [69].

Participants’ recommendations directly related to this lack of privacy or an appropriate space; they advocated for “a warm and comfortable space” to be made available [71].

> “Perhaps there should be some rule where they could sleep together once a week or something like that in a flat or in a room or in their room. They shouldn’t have to hide”[67].

#### Lack of opportunity to form and maintain relationships: service environment

The prohibition of sexual relations by staff and the possibility of making others uncomfortable deterred service users from being intimate within inpatient mental health services [30,36,63]. This extended to a broader inability to participate in the ‘normal activities’ that couples typically engage in, which was seen as not conducive to maintaining a romantic relationship:

> “You’re very limited as to what you can do here. It’s not like you can go to the movies or go and see a band or anything like that… Most people in a relationship get to have sex in the night time. This can never happen for us” [67].

An absence of opportunity “to mix and meet people” was also highlighted by participants, with funding cuts at day centres being listed as a contributor to this issue [61]. While the majority of participants were eager to form and maintain relationships [36], one found that merely being in the service environment (an inpatient hospital with an outpatient clinic) left them apathetic toward relationships in general:

> “Suddenly locked up in this closed environment I am not connected to anything personally or spiritually I am just kind of there” [71].

### Lack of support from staff and services

Individuals reported that services and staff neglected to acknowledge their romantic and intimate relationship needs [30,61–63,69,70].

They expressed that services’ “biochemical” [69] and medical-model stance resulted in unsatisfactory handling of relationship-based queries:

> “Sometimes [mental health practitioners] give you the usual textbook answers in terms that are too specific to be understood properly. Moreover they judge it as frivolous and unimportant and this hurts a lot” [63].

The delivery of care under this approach meant that “appointments are typically brief intermittent and focused on symptoms rather than psychosocial issues – [which] was a barrier to effective therapeutic relationships” [70]. Additionally, participants noted that staff were stretched too thin [62,65] making them “harder to reach” [65], and others felt “that they’re generally not approachable” [30]. Some service users felt that the staff’s unavailability was “because nobody cares” [62].

In instances where sexual activity was discussed with professionals, participants were often met with awkwardness [69] or felt they were “treated like children” [66]. One participant [69] revealed that after disclosing a sexual harassment incident, “nothing happened.”

### Recommendations for communication within services on romance and intimacy

Service users had proposals to improve the provision of romance and intimacy-related support. Four studies recommended that mental health practitioners display an openness to discussing the topic [13,30,63,66]. Doing so was seen as a first-step toward meeting support needs, as well as critically recognising “that there is a romantic aspect to a patient as well… [they’re] a human being” [70]. In opening-up discourse surrounding intimacy, it was noted that clinicians required training in appropriate, sensitive [68] and supportive communication [66]. This necessitated professionals possessing an ability to be reciprocal to the client:

> “Be sensitive to whether the person reacts by feeling they want to open up and discuss or wants to clam up shut down and say nothing and respect that” [70].

Creating support groups was also recommended [62,70]. However, one participant [62] disagreed:

> “It’s intimate you don’t get into it in group sessions.”

### Desire for advice and education on sexual intimacy, romantic relationships, and safe sex

Overall, studies advocated for advice and education within services to address a scarcity of resources regarding romantic/intimate relationships [13,62,63,71]. These recommendations were drawn from authors’ interpretations of participant’s recognition of the lack of information, rather than direct recommendations from service users themselves [13,62,63,71].

> “To say we received sexual education—no not really… it was never discussed”[71].

At the prospect of services introducing resources related to sexual health and measures for safe sex, participants had diverging views. Some displaying enthusiasm [13] and others did not see it as a priority [63,68].

## Discussion

The aim of this study was to explore romantic and intimate relationship needs of mental health service users and establish their self-identified support needs. We found that service users generally wanted romantic and intimate relationships, which is consistent with the findings of previous qualitative [33,48,49] and quantitative reviews [10]. In keeping with previous research, participants acknowledged the potential benefits of romance and intimacy - such as improved prognosis, sense of self-worth, and emotional regulation [11,26,67]. However, having a mental health condition presented specific challenges in forming and maintaining romantic/intimate relationships. These barriers also concurred with the findings of previous reviews that had focused more on sexual experiences rather than romantic and intimate relationships, including personal, interpersonal, and social factors [33,48,49]. We also identified service level barriers, in keeping with past research [26,48].

These included negative attitudes from clinical staff about service users being able to have romantic/intimate relationships, lack of resources to support them, and environmental factors, such as a lack of privacy.

Findings from the included studies consistently emphasised that having a mental illness impacted negatively on people’s ability to pursue a romantic/intimate relationship, though this was quite a nuanced point, with the specific issues being highly individual. This concurs with McCann’s [48] recognition of the “complexity of individual experiences” in this area. Previous studies have outlined the relevance of how psychological, emotional, sociological, and pharmacological factors can each interfere with people’s ability to meet their own romance/intimacy needs [34,38,72]. This complexity amongst individuals’ experiences should be kept in mind when considering results.

The impact of mental illness influenced some participants’ self-perception, negatively affecting their ability and desire to form relationships. These perceptions often originated from the impact of their symptoms, leading to some feeling incapable of managing these alongside a relationship. Social isolation and lack of social confidence also led to people feeling unable to initiate conversations with others, a finding also highlighted by Brand et al. [33]. Further, experiences of discrimination and stigma exacerbated people’s internalised stigma [38], with some participants feeling that they were unfit to have a relationship [59,64,71]. This review also identified that difficulties initiating relationships were exacerbated by fears of rejection, pre-emptive guilt and concerns about burdening a potential partner due to their mental health problems, as well as past experiences of sexual abuse [27,28,48]. These kinds of experiences and perspectives aligned with McCann’s and Hortal-Mas et al.’s findings [48,49]. McCann noted “difficulty in acceptance of self and feelings of inadequacy” [59,64,71] while Hortal-Mas et al. described how people gradually let go of wanting a relationship over time [49]. Similarly, a sense of resignation about not being able to have a relationship, or of not wanting to have one due to a need for ‘self- prioritisation’ emerged from service users’ perspectives.

The phenomenon of ‘learned helplessness’ [73] may shed light on how individuals gradually became resigned to not having a relationship [74]. Symptoms, discrimination and stigma [38], social isolation, and perceived lack of support may have contributed to some believing that efforts to form relationships would inevitably fail, and they thus avoided initiating them. However, in contrast, many service users did remain hopeful about forming a relationship in the future. These dispersed accounts may have been indicative of psychological resilience [75], as a mechanism of coping with the challenges faced [64,76].

Through discussing and understanding the obstacles service users faced in meeting their needs, mental health practitioners may be better equipped to respond to barriers and tailor supports. As it stands, the results emphasised that staff generally do not talk about romance/intimacy [33,48,77]. Service users felt that this stemmed from clinical attitudes that were risk averse, concurring with Brand et al. [26]. On the other hand, participants displayed an interest in receiving service support in relation to romance/intimacy. They expressed a desire to talk about this topic with staff and thought that doing so would be affirming, constructive, and informative, as also highlighted by McCann [48]. Importantly, there was a sense that recognising service users’ romance/intimacy needs would contribute to feeling seen as a ‘whole-person’ rather than through a clinical lens [63,69].

Mental health staff’s avoidance of discussing romance/intimacy, undermines the recommendations that mental health services should be person-centred. Specifically, it represents an omission in offering a recovery-orientated approach, which emphasises holistic, personalised, and collaborative support to empower people’s autonomy and dignity [26,78]. This lack of focus on relationships may deprive service users of intimacy, affection and support that could promote their recovery, and lead to loneliness, depression, and stress [79].

## Service implications

Previous qualitative studies with staff participants suggest that the lack of support from services may result from staff feeling ill-equipped to address romantic and intimate needs [21,24] coupled with a belief among staff and service users that this topic remains taboo [20,37]. The apparent avoidance of this topic may be compounded by concerns about service users’ sexual vulnerability and risk of exploitation, either currently or historically.

Such concerns raise key questions relating to ethics and safeguarding [31,42]. Perhaps unsurprisingly, these findings have been discussed more in the context of higher risk inpatient services. Nonetheless, safeguarding guidelines within services should feature a co-creation approach that considers the potential for exploitation [27,28] alongside service users’ capacity and hopes for relationships [66]. English national guidelines recognise that only through discussing people’s romantic and sexual activity can staff offer support for maintaining sexual safety and developing healthy relationships [17], as endorsed by the findings of this review.

The findings related to previous sexual abuse experiences, in-part reinforced staffs’ concerns for vulnerability [27,28,30], but also affirm the growing focus on trauma-informed care within services [80,81]. Acknowledgment that any service user may have had experiences of exploitation signifies the importance of being sensitive to past trauma when providing care in general, including when discussing relationships. Relationships can be negatively impacted by previous experiences of sexual abuse [82], and an inappropriate response from services to disclosure of such experiences may exacerbate the associated trauma [69,77]. There is thus a need for services to train staff in addressing such disclosures appropriately, and to develop guidance on how staff can provide individualised support to service users with regard to building safe and loving intimate relationships (21,28,29,43). In the event of a disclosure, offering a space to explore and/or provide support for previous experiences should be offered or signposted [70].

Inpatient and supported accommodation services also need to consider how to provide environments where service users can potentially engage in consensual intimate/sexual relationships in privacy and with adequate safeguards in place [32,34,35]. Additionally, the effects of mental illness on service users’ partners must be recognised, and practitioners made aware of the potential stresses for partners. An option of involving partners in treatment should be considered, where feasible, echoing previous recommendations [45,48]. Clearly, specialist training and resources for staff to support service users and partners alike in this area is urgently needed so that the inclusion of romantic and intimacy needs can be incorporated into care plans.

Clinicians should be encouraged to make patients aware that discourse around hopes and needs for romantic/intimate relationships is permitted. In acknowledging these needs, open and receptive communication is particularly important. Design of new initiatives should encompass informing individuals of their rights to relationships and promoting the development of healthy relationships [23,33], regular medication reviews to minimise sexual side effects [26], promote self-esteem and address dealing with stigma, facilitate optional support groups, and create social activities as a means of meeting others and developing social skills [9,33,45]. This review emphasised that building trusting service user-professional relationships is a crucial step toward a successful provision of these strategies.

While some of the recommendations of this review pertained more specifically to hospital services, the implications likely extend to community-sector organisations, local authority services, and charities. Irrespective of service type, recognising relationships needs and incorporating service user-oriented support measures into care was tied with improving prognoses [7,9] and satisfaction with services [5,8,67]. Moreover, the impact of mental health symptoms and medication side effects is often less pronounced in community settings, potentially increasing the importance of supporting romance and intimacy needs. Community settings also offer enhanced opportunity to establish enduring therapeutic relationships more conducive to discussing these topics.

## Strengths and limitations of this review

Articulating the voices of service users was a strength and priority of this research. While this review shared commonalities with the qualitative systematic review conducted by McCann et al. [48], it served as an important update given the growing interest on this topic. This review extended beyond the experiences of sexuality amongst those with severe mental health problems and, including a broader focus on the needs for romantic/intimate relationships for people with any mental health condition.

The included papers were all rated as having high methodological quality, providing greater confidence in the results. Furthermore, findings from each study were mostly rated as unequivocal, largely due to the alignment between authors’ findings and participant quotes (illustrations).

However, while the JBI critical appraisal tool is well-established and the GRADE-CERQual is recommended by the JBI, ratings remained open to a certain degree of subjective judgement, especially given that these were conducted by one rater.

Our findings established areas of difficulty for service users, often without explicitly defining the support required to overcome them. While providing some useful insights to inform service improvements, data tended to emphasise barriers rather than solutions as to how individuals best wished to be supported. In addition, while the meta-aggregate approach acted to minimise subjective interpretations by retaining the standpoint of the included studies’ authors, it remained a synthesis of those authors’ interpretations.

Moreover, our study team did not include any people with lived experience of mental illness, so this important perspective did not inform our analysis or interpretation of the results.

However, the consistency of findings across studies and their congruence with most of the existing literature on the topic provides confidence in our findings.

## Limitations of the primary papers

The included studies mainly focused on people with serious mental illness, which may contribute to the similar results from this and previous reviews [47,48]. Consequently, the objective of gathering a perspective that may be applicable to other mental health service users was not achieved [83]. Our findings may therefore not all be relevant to community-based services, especially those working with people with common mental disorders rather than serious mental illness. Most studies being from high-income countries (14/17) may also limit the generalisability of findings to other settings. Additionally, lived-experience involvement was minimal among studies (4/17), potentially limiting the relevance and applicability of recommendations to real-world contexts and the actual preferences of service users. Finally, at times, individual study’s findings lacked clarity in their wording, however in following JBI guidelines, verbatim extractions were not edited.

## Future research

Future research would benefit from incorporating lived-experience into the study process, which may help to refine research questions and guide understanding of potential supports. For example, this paper’s research question, which focused entirely on needs for romantic/intimate relationships, failed to acknowledge the absence of need discovered in some papers [59,63–65,71].

Further qualitative studies are needed to explore experiences of service users without serious mental illness and those living in community settings. Specifically, future research is needed to assess potentially differing views within different mental health care settings. The focus on risk-aversion and containment within services may be reflective of the scarcity of research from community settings – which future research should address.

Further research from the voluntary sector may be particularly useful, as such services may reach parts of the community that have poor trust in and engagement with statutory services, and often provide holistic, relational care [84]. Additionally, research is needed to assess whether populations with non-serious mental illness would still value romance/intimacy-related support from services and examine their views romance/intimacy needs.

Given the small number of studies identified from outside high-income countries, there is a need to conduct research in less economically developed countries. It is also important to examine service user populations within specific cultural groups, where attitudes towards relationship support seeking behaviours may vary [85]. Given that clinicians’ attitudes and cultural barriers were noted in Raisi et al.’s [68] and Yang et al.’s [71] studies, further research should consider perspectives based on different cultural contexts. Of the included papers, these were the only two performed in Eastern populations (China and Iran).

In addition, research studies are required that investigate how to best enable people with mental health conditions to challenge their internalised stigma, and to develop and test interventions that can help build confidence to establish and maintain interpersonal relationships. Further research should consider the utility of interventions that address the barriers to romantic and intimate relationships – such as fear of rejection, past sexual abuse, and partner burden-related guilt. It is important for research and intervention design to take a trauma-informed approached as this topic can be highly sensitive [80]. Additionally, the potential utility that support groups may have, and for whom, could be established through subsequent qualitative research.

With future intervention development in mind, further research should explore whether designing a structured intervention to assist with romantic and intimate relationship needs is achievable, given the personal nature and complexity of the subject. Nevertheless, meaningful conversations with practitioners and providing tailored support based on individual needs are crucial steps towards improving outcomes.

## Conclusion

Mental health service users expressed a desire for romantic and intimate relationships in much the same way that those without mental health conditions often do [86]. However, there were factors related to mental illness that hindered attaining romantic and intimate relationships on a personal, interpersonal, social, and service level. In general, service users were not receiving assistance from mental health services to find intimate/romantic relationships due to specific barriers. More research is required to inform the training and guidance staff require to be able to approach this topic in their discussions with service users and incorporate specific actions into tailored care plans.

## Supporting information

S1 Table. PRISMA Checklist.

S2 Table. Search strategies for each data base.

S3 Table. JBI Critical Appraisal Checklist for Qualitative Research.

S4 Table. Findings and illustrations from each paper.

S5 Table. Details of aggregating findings and illustrations into categories.

S6 File. GRADE-CERQual ratings.

S7 File. PROSPERO Protocol.

## Data Availability

All data produced in the present work are contained in the manuscript.

## Acknowledgments

I would like to express my sincere gratitude to my supervisors, Brynmor Lloyd Evans, Helen Killaspy, and Sharon Eager, for their invaluable guidance and unwavering support throughout this research project. Their expertise and insights were critical to the foundations and completion of this project. I extend my genuine thanks to Eunice Wu, without whom the systematic review process would not have been possible.

## Supporting information

S1 Table. PRISMA Checklist.

S2 Table. Search strategies for each data base.

S3 Table. JBI Critical Appraisal Checklist for Qualitative Research.

S4 Table. Findings and illustrations from each paper.

S5 Table. Details of aggregating findings and illustrations into categories.

S6 File. GRADE-CERQual ratings.

S7 File. PROSPERO Protocol.

