## Supplementary material for "Systematic review and meta-aggregate analysis of mental health service users’ perspectives on romantic and intimate relationships and their support needs": S2 Table. Search strategies for each data base.

Search strategies for each data base

***MEDLINE***

| **#** | **Query** | **Results from 9 April 2024** |
| --- | --- | --- |
| 1 | exp mental disorders/ | 1,483,787 |
| 2 | exp mental health/ | 66,619 |
| 3 | ("romantic relationship*" or romance or love or courtship or dating or "romantic partners" or "romantic involvement" or "romantic attachment").tw. | 34,614 |
| 4 | ("intimate relationship*" or intimacy or "emotional connection" or "close relationship*" or "romantic intimacy").tw. | 30,750 |
| 5 | ("sexual behavio*" or "sexual activit*" or "sexual relationship*" or sexuality or "sexual health" or "sexual practice*").tw. | 68,878 |
| 6 | (view* or opinion* or perception* or perceived or attitude* or knowledge or experience*).tw. | 3,306,488 |
| 7 | (qualitative or ethnography or phenomenology or "grounded theory" or interview* or "focus group*" or "content analysis" or survey*).tw. | 1,515,393 |
| 8 | (service user* or patient or client or consumer or "people nr service*").mp. [mp=title, book title, abstract, original title, name of substance word, subject heading word, floating sub-heading word, keyword heading word, organism supplementary concept word, protocol supplementary concept word, rare disease supplementary concept word, unique identifier, synonyms, population supplementary concept word, anatomy supplementary concept word] | 3,508,050 |
| 9 | 1 or 2 | 1,528,845 |
| 10 | 3 or 4 or 5 | 129,130 |
| 11 | 6 or 7 | 4,234,303 |
| 12 | 8 and 9 and 10 and 11 | 1,297 |

***PsycINFO***

| **#** | **Query** | **Results from 9 April 2024** |
| --- | --- | --- |
| 1 | exp mental disorders/ | 1,105,802 |
| 2 | exp mental health/ | 97,039 |
| 3 | ("romantic relationship*" or romance or love or courtship or dating or "romantic partners" or "romantic involvement" or "romantic attachment").tw. | 55,464 |
| 4 | ("intimate relationship*" or intimacy or "emotional connection" or "close relationship*" or "romantic intimacy").tw. | 30,312 |
| 5 | ("sexual behavio*" or "sexual activit*" or "sexual relationship*" or sexuality or "sexual health" or "sexual practice*").tw. | 71,598 |
| 6 | (view* or opinion* or perception* or perceived or attitude* or knowledge or experience* or person-cent*).tw. | 1,876,183 |
| 7 | (qualitative or ethnography or phenomenology or "grounded theory" or interview* or "focus group*" or "content analysis" or survey*).tw. | 889,650 |
| 8 | (service user* or patient or client or consumer or "people nr service*").mp. [mp=title, abstract, heading word, table of contents, key concepts, original title, tests & measures, mesh word] | 534,706 |
| 9 | 1 or 2 | 1,169,992 |
| 10 | 3 or 4 or 5 | 144,047 |
| 11 | 6 or 7 | 2,271,780 |
| 12 | 8 and 9 and 10 and 11 | 1,331 |

***CINAHL***

| **#** | **Query** | **Results from 9 April 2024** |
| --- | --- | --- |
| S5 | S1 AND S2 AND S3 AND S4 | 430 |
| S4 | AB (service user* OR patient OR client OR consumer) | 1,769,870 |
| S3 | AB ( view* OR opinion* OR perception* OR perceived OR attitude* OR knowledge OR experience* ) OR AB ( qualitative OR ethnography OR phenomenology OR "grounded theory" OR interview* OR "focus group*" OR "content analysis" OR survey*) | 1,072,184 |
| S2 | AB ( "romantic relationship*" OR romance OR love OR courtship OR dating OR "romantic partners" OR "romantic involvement" OR "romantic attachment" ) OR AB ( "intimate relationship*" OR intimacy OR "emotional connection" OR "close relationship*" OR "romantic intimacy" ) OR AB ( "sexual behavio*" OR "sexual activit*" OR "sexual relationship*" OR sexuality OR "sexual health" OR "sexual practice*" ) | 42,988 |
| S1 | AB ("mental health" OR "mental illness" OR "mental disorder" OR "psychological disorder" OR "psychiatric disorder" OR "psychological distress" OR "psychiatric illness" OR "psychological problem*" OR "psychiatric problem*" OR "mental health issue*" OR "psychological issue*" OR "psychiatric issue*" OR "mental health condition*" OR "psychological condition*" OR "psychiatric condition*" OR "mental health challenge*" OR "psychological challenge*" OR "psychiatric challenge*") | 143,071 |

***Web of Science***

| **#** | **Query** | **Results from 8 April 2024** |
| --- | --- | --- |
| 1 | TS=(mental health OR mental disorder*) | 521,144 |
| 2 | AB=("romantic relationship*" OR romance OR love OR courtship OR dating OR "romantic partners" OR "romantic involvement" OR "romantic attachment") | 772,754 |
| 3 | AB=("intimate relationship*" OR intimacy OR "emotional connection" OR "close relationship*" OR "romantic intimacy") | 55,866 |
| 4 | AB=("sexual behavio*" OR "sexual activit*" OR "sexual relationship*" OR sexuality OR "sexual health" OR "sexual practice*") | 78,753 |
| 5 | AB=(view* OR opinion* OR perception* OR perceived OR attitude* OR knowledge OR experience*) | 5,971,991 |
| 6 | AB=(qualitative OR ethnography OR phenomenology OR "grounded theory" OR interview* OR "focus group*" OR "content analysis" OR survey*) | 1,396,982 |
| 7 | AB=(service user* OR patient OR client OR consumer) | 6,999,043 |
| 8 | #2 OR #3 OR # | 895,987 |
| 9 | #5 OR #6 | 6,752,739 |
| 10 | #1 AND #7 AND #8 AND #9 | 2,697 |
