## Supplementary material for "Systematic review and meta-aggregate analysis of mental health service users’ perspectives on romantic and intimate relationships and their support needs": S3 Table. JBI Critical Appraisal Checklist for Qualitative Research.

JBI Critical Appraisal Checklist for Qualitative Research

| **References** | **Q1** | **Q2** | **Q3** | **Q4** | **Q5** | **Q6** | **Q7** | **Q8** | **Q9** | **Q10** | **Score** | **Quality** |
| --- | --- | --- | --- | --- | --- | --- | --- | --- | --- | --- | --- | --- |
| Brand et al. (2022) | Y | Y | Y | Y | Y | N | N | Y | Y | Y | 8/10 | High |
| Brown et al. (2014) | Y | Y | Y | Y | Y | Un | Y | Y | Y | Y | 9/10 | High |
| Budziszewska et al. (2020) | Y | Y | Y | Y | Y | Y | Y | Y | Y | Y | 10/10 | High |
| deJager et al. (2017) | Y | Y | Y | Y | Y | N | N | Y | N | Y | 7/10 | High |
| Forrester-Jones et al. (2023) | Y | Y | Y | Y | Y | N | Y | Y | Y | Y | 9/10 | High |
| Kaplan et al. (2022) | Y | Y | Y | Y | Y | Y | Y | Y | N | Y | 9/10 | High |
| Landi et al. (2020) | Y | Y | Y | Y | Y | N | Y | Y | Y | Y | 9/10 | High |
| McCann (2010) | Un | Y | Y | Y | Y | N | N | Y | Y | Y | 7/10 | High |
| Mizock et al. (2020) | Y | Y | Y | Y | Y | N | Y | Y | Y | Y | 9/10 | High |
| Ostman (2016) | Y | Y | Y | Y | Y | Y | Y | Y | Y | Y | 10/10 | High |
| Quinn and Happell (2016) | Y | Y | Y | Y | Y | N | N | Y | Y | Y | 8/10 | High |
| Quinn and Happell (2015a) | Y | Y | Y | Y | Y | N | N | Y | Y | Y | 8/10 | High |
| Quinn and Happell (2015b) | Y | Y | Y | Y | Y | N | N | Y | Y | Y | 8/10 | High |
| Raisi et al. (2017) | Y | Y | Y | Y | Y | N | N | Y | Y | Y | 8/10 | High |
| Robertson et al. (2015) | Y | Y | Y | Y | Y | N | Y | Y | Y | Y | 9/10 | High |
| White et al. (2021) | Y | Y | Y | Y | Y | N | Y | Y | Y | Y | 9/10 | High |
| Yang et al. (2023) | Y | Y | Y | Y | Y | N | N | Y | Y | Y | 8/10 | High |

Criteria met: Yes (Y), No (N), Unsure (Un)

Critical appraisal questions for comparable qualitative studies:

1. Is there congruity between the stated philosophical perspective and the research methodology?

2. Is there congruity between the research methodology and the research question or objectives?

3. Is there congruity between the research methodology and the methods used to collect data?

4. Is there congruity between the research methodology and the representation and analysis of data?

5. Is there congruity between the research methodology and the interpretation of results?

6. Is there a statement locating the researcher culturally or theoretically?

7. Is the influence of the researcher on the research, and vice- versa, addressed?

8. Are participants, and their voices, adequately represented?

9. Is the research ethical according to current criteria or, for recent studies, and is there evidence of ethical approval by an appropriate body?

10. Do the conclusions drawn in the research report flow from the analysis, or interpretation, of the data?
