## Supplementary material for "Systematic review and meta-aggregate analysis of mental health service users’ perspectives on romantic and intimate relationships and their support needs": S5 Table. Details of aggregating findings and illustrations into categories.

Details of aggregating findings and illustrations into categories

**Service users have a desire for romantic and intimate relationships: the normalcy, unmet need, and complexity of fulfilling these hopes**

*Normality of sexual and intimate relationships and its perceived benefit*

| **Findings and citations** | **Illustrations** | **Level of credibility** |
| --- | --- | --- |
| Sex as normal human behaviour  *Quinn & Happell, 2015b* | *“It's quite normal it's something everyone does. It doesn't matter where you find your partner. You can't pick where and who you fall in love.”* | Unequivocal |
| Having a sexual relationship as normal human behaviour  *Quinn & Happell, 2015a* | *“The staff have got their relationships people in the community have relationships the security officers the policemen prison officers they all have relationships. Just because we draw a pension and we’re under the Mental Health Act doesn’t mean that we don’t have needs and wants for a relationship and sex.”* | Unequivocal |
| The feeling of being loved are therapeutic and an aspect of their lives that supports their personal recovery  *Quinn & Happell, 2015a* | *“I reckon it’d be very therapeutic. The sense of feeling loved by another person can help you progress quicker and supporting each other through stuff that nurses can’t help you with. It helps you with certain emotions like if you are a male and you’re a bit aggressive it can help you mellow out.”* | Unequivocal |
| Intimate relationships as beneficial to recovery  *Robertson et al., 2015* | *Intimacy was experienced in relationships where participants felt accepted supported and understood. These factors were beneficial to recovery and were especially important for participants who felt marginalized and misunderstood on the basis of their sexual identity and mental health difficulties.* | Unequivocal |
| Intimate relationships allowed participants to feel “normal” and access a sense of social inclusion  *Robertson et al., 2015* | *“She did come and see me and we went out to the park and um we went for walks and things like that we went for something to eat um … (later on in transcript) it just felt like I had some normal a bit of that it wasn’t the normal … the normality.”* | Unequivocal |
| Conceptualising romantic relationships: impact on wellbeing  *White et al., 2021* | *“I think romantic relationships are incredibly important for anyone regardless of whether you have a mental health problem or not I think that romantic relationships make you feel better about yourself they give you a sense of self-worth a sense of identity a sense of self purpose.”*  *“… just noticed she didn’t really feel the same way about me as I felt the same about her [ …] it damages your confidence definitely.”* | Unequivocal |
| Sex is an effective way to regulate emotion  *Yang et al., 2023* | *“I always thought and believed that such intense and happy sexual experiences would make me calmer I had always had a good sexual relationship with my husband when my sexual function was not so bad and I feel much happier as a person.”* | Unequivocal |

*Unfulfilled desire for romantic and intimate relationships*

| **Findings and citations** | **Illustrations** | **Level of credibility** |
| --- | --- | --- |
| Participants were not sexually active, but eager to be in a sexual relationship  *Brand et al., 2022* | *“I don't have anyone I am on my own… I do still like to get married one day.”*  *“I want to be in a stable relationship but I am not really trying at the moment …”* | Credible |
| Amputation: entry to the medium-secure unit comes at cost of losing one’s sexuality  *Brown et al., 2014* | *“I think it (sexuality) is relevant because I think it – I associate it with just general vibrancy … And – life force and a dynamic that’s gone missing it’s disappeared.”* | Unequivocal |
| Affective needs of sexual intimacy  *Landi et al., 2020* | *“For me sex with the partner has always been the most important thing in life. I would like to have a woman a girlfriend.”* | Unequivocal |
| Emotional sorrow: lack of human relationship  *Landi et al., 2020* | *“I had a life so scarce of human relationships that now I consider precious any kind of feeling expressed to me. Even when someone is disappointed by me actually I am glad because I can go home saying: 'at least today I had a human contact!'”* | Unequivocal |

*The desire for and complexities of parenthood*

| **Findings and citations** | **Illustrations** | **Level of credibility** |
| --- | --- | --- |
| Family planning and parenting  *McCann, 2010* | *“I’d really like to have children but maybe it’s too late now. We’re trapped in this place.”*  *“At the moment because of my illness – in case I get sick and they get put into a home… no children.”* | Unequivocal |
| Pressure to take the birth control pill  *Landi et al., 2020* | *“Regarding contraception healthcare professionals agreed with my parents that it was better for me to take the pill. But I did not want to I would love another pregnancy.”* | Unequivocal |
| Having children – rewarding but risky  *White et al., 2021* | *“You know love connection and many other things having children I mean I dunno if I’ll be able I hope to be able to have children […] it’s very important to me.”*  *“I was always a little bit concerned that I could have a puerperal psychosis relapse and then you’re in a nightmare situation of your children taken into care and stuff like that could happen or my mother would’ve had to step in […] would I have coped with a career and children and a mental illness and a husband who may or may not have understood? I don’t know […] you potentially could harm your own children if you weren’t well I know of a case where that happened that would be devastating.”* | Unequivocal |

**Mental health conditions directly hinder romantic and/or intimate relationships at the individual-level**

*Impact of Medication’s Side Effects*

|  | **Findings and citations** | **Illustrations** | **Level of credibility** |
| --- | --- | --- | --- |
|  | Medications as barriers  *Brand et al., 2022* | *While some participants recognized and acknowledged the positive effects medications have on their mental state, they also recognized this came with a detriment to their sexual function.* | Unequivocal |
|  | Symptoms and side effects of medication  *deJager et al., 2017* | *“It’s not just because of the medication that you can’t get an erection, it is also because of performance anxiety. To be unable to get an erection affects your whole sense of being a man.”* | Unequivocal |
|  | Mental health symptoms and side-effects of medications  *Forrester-Jones et al., 2023* | *“The current medicines I’m on, the side effects mean I wouldn’t really be much use [in a relationship] so I might have to get yet more medicine to help counteract that. I mean I’m quiet shy at the moment, I’m quite depressed and that’s not a particularly attractive quality. You don’t want to be looking for a relationship to solve all your problems, but I personally think it’s part of a healthy life. I think just the pattern of my life. The lack of stability, not having a job and things like that, not being in mainstream society.”* | Unequivocal |
|  | Medication side effects  *Kaplan et al., 2022* | *“I want to have sex 3–4 times, and I can't because I don't have a good erection.”* | Unequivocal |
|  | The effects of medication upon their sex life  *McCann, 2010* | *“He [the doctor] put me on an antidepressant and I went right off sex altogether. I think that is probably one of the reasons we broke up... I think it is all these drugs that, affected, what do they call it. You’re sex drive or something.”*  *“I began to have lots of sex and I wanted more and more and I was so horny on Prozac. My sex drive went super high, I was stuck to the ceiling, so high it was incredible.”* | Unequivocal |
|  | Sexual dysfunction as a result of psychoactive medication  *Robertson et al., 2015* | *“I’m off the medication now, it wasn't the only reason but it was one of the reasons because it, my poor partner, then partner, would be ex- (laughs), ex-, at the point of exhaustion because it would take, take so long [to achieve orgasm].”* | Unequivocal |
|  | Side effects from antipsychotic medication  *White et al., 2021* | *“My medication causes me to eat more and I put on weight, which has knocked my confidence a bit.”* | Unequivocal |
|  | Side effects of antipsychotics  *Yang et al., 2023* | *“When I have sex with my lover, sometimes I cannot get an erection. Even if I have an erection, it does not last for a while. That really sucks.”* | Unequivocal |

*Impact of symptoms*

| **Findings and citations** | **Illustration** | **Level of credibility** |
| --- | --- | --- |
| Illness-induced changes in experiencing love can be obstacles to entering and maintaining romantic relationships  *Budziszewska et al., 2020* | *“For me that’s a very intimate form of bond. If you hear someone’s voice in your head and that voice is with you non-stop then it’s like no one else can become closer to you than that voice in your head.”* | Unequivocal |
| Specific sexual problems from mental illness  *McCann 2010* | *“Well it takes a long time and there’s that feeling of suffocation. I mean sexually and emotionally. I have a really strong psychological hold over my sort of sexual drives and feelings. I am sure that is why I have never had an orgasm because there is something in me that always holds back and always sets apart and removes myself from the sexual act.”* | Unequivocal |
| Symptom interference with the development and maintenance of a relationship  *Mizock et al., 2019* | *“It was very difficult for me to date because I was getting sick all the time. So it was very hard for me to concentrate and really get to work on finding somebody … I was sick I was sick. I just remember I was sick. It’s only now that I’m not so bipolar.”* | Unequivocal |
| Psychotic symptoms: barrier hindering sexual activity  *Yang et al., 2023* | *“I sometimes have an erection in the morning but I do not care. Because at the moment I am out of control I’m not too…I’m not very trusting in myself to do anything else.”* | Unequivocal |

*Self-prioritisation: romantic and intimate relationships as secondary to mental helath condition*

| **Findings and citations** | **Illustrations** | **Level of credibility** |
| --- | --- | --- |
| Illness adjustment is all-consuming at first and can lead to isolation  *Budziszewska et al. 2020* | *“The beginning of this illness is that you don’t know what you want where you stand and who you are. You have millions of questions in your head so you just forget about stuff like love. It’s set on the back burner right? Only after a certain period of time when you pull yourself together you start thinking about stuff like that right? The first stage is ME only.”* | Unequivocal |
| Voluntary sexual abstinence  *Budziszewska et al. 2020* | *“But I just feel that I’m not ready and I can’t imagine being ready. So it’s a little easier for me this way.”* | Unequivocal |
| Affective needs: antithetic positions  *Landi et al., 2020* | *“I do not feel the need [for sexuality] I do not know why but actually do not feel it.”* | Unequivocal |
| Giving up on sexual intimacy  *Mizock et al., 2019* | *“What dating life? I’ve never had a dating life. I don’t have a dating life. I’m too busy trying to be — it’s all I can do to take care of myself. I can’t do relationships … I think also … I put a huge amount of focus into my mental health all the time.”* | Unequivocal |
| Sexual relationships secondary in the case of serious mental illness  *Ostman 2014* | *“Every day is a struggle and there is no place for sexuality. Since the illness began I have prioritized keeping the anxiety away from me and wrestling with my work. The main thing is I have to be strong. That’s why I can’t prioritize sex life.”* | Unequivocal |
| Stop meeting their own sexual needs to focus on controlling the symptoms of their mental disorder  *Yang et al., 2023* | *“I have prioritized keeping the schizophrenia away from me and wrestling with my work. The main thing is I have to be strong. That’s why I cannot prioritize sex life.”* | Unequivocal |

**Psychosocial factors impact romantic and/or intimate relationships**

*Social difficulties to forming and maintaining relationships as a consequence of mental illness*

| **Findings and citations** | **Illustrations** | **Level of credibility** |
| --- | --- | --- |
| Difficulties in socializing and communication  *Brand et al., 2022* | *“I just stay home most of the time; don't get out… I think it's because of my illness.”* | Unequivocal |
| Lack of social skills and experience  *deJager et al., 2017* | *“I can’t pull it off. I’m my worst enemy … (…) When I encounter people I have no idea what topics are interesting or if he/she will like what I have to tell him/her. Nerves.”* | Unequivocal |
| Challenges in forming and maintaining relationships: meeting people who are incompatible  *McCann 2010* | *“I look at girls in the street but don’t know what to do. The kind of girl’s I meet in the drop-in . . . they are having a hard time in their heads. That’s not the kind of girl I want. I want one I can marry and have kids with. I met a girl in the hospital and we saw each other for a while . . . Wanted to have children and marry but it’s that stigma thing . . . People with mental illness only meet the same type of people with the same type of problems. It’s hard man.”*  *“I am seeing a woman in the hostel. We have been together for three years. She has schizophrenia and learning difficulties. We want to get married and live together. The staff have only found out that we are seeing each other and are not happy.”* | Credible |
| The challenge women with serious mental illness may face in finding a partner at a similar level of mental health functioning  *Mizock et al., 2019* | *“I don’t want to live a life where there is the possibility of instability. Because I consider myself very stable right now … I had some dates with this guy that I found out spent 2.5 years in [name redacted] State Hospital and 18 years in a group home. He has only been in his own place for 8 months now. I had grown so much more than he had. I had to drag him along …”* | Unequivocal |

*Discrimination and (self-)stigma*

| **Findings and citations** | **Illustrations** | **Level of credibility** |
| --- | --- | --- |
| Low status and discrimination  *Budziszewska et al. 2020* | *“Unfortunately he found out I’m in [name of major psychiatric hospital] and he didn’t want to be with me anymore.”* | Unequivocal |
| (Self-)stigma  *deJager et al., 2017* | *“I was thinking: you will lose him for sure. Who would want someone with a psychosis?”* | Unequivocal |
| Self-stigma  *Forrester-Jones et al. 2023* | *“If I were to meet somebody I’d prefer that they weren’t mentally ill. And I wouldn’t like to have a relationship with another schizophrenic. But I’d like to meet somebody who would take me out of that – a normal person – a person who doesn’t suffer with mental health problems so I could live their way instead of worrying and anxiety.”* | Unequivocal |
| Social stigma  *Forrester-Jones et al., 2023* | *“And I wouldn’t tell them that I was schizophrenic – I’d say I was paranoid because I wouldn’t want them looking at me funny to start with and thinking “oh is she going to attack me?”* | Credible |
| Stigma and self-esteem  *McCann 2010* | *“I am reluctant [to approach women] because I’m afraid they all know that I am not well. I am very reluctant to go next to my own Kurdish people because of the shame I feel.”*  *“The only thing that would stop me [meeting someone] is that I was neglecting myself.”* | Credible |
| Dating barriers due to stigma  *Mizock et al., 2019* | *“I would like to meet somebody but you know who would want me? I don’t work. I’m a bipolar. Who would want to get involved with me?… I would like to meet someone and get married again. I’ve dated a few times — three different guys — and it’s strange how you forget how to behave … No one would want to be involved because they would think that I was not stable. They would have the problem with the stigma.”* | Unequivocal |
| Prejudice and discrimination as barriers to service users forming and maintaining intimate relationships  *Robertson et al., 2015* | *“[The psychiatrist said] that my my my problems were emotional and that being lesbian or gay was was a contributory factor to my to my mental health.”*  *“Staff welcomed X [partner] onto the ward and were happy to see him and so it was easy for him to come in um made it more pleasant for him to be there um that … meant he was quite happy to come as much as he wanted to come and see me and that was obviously good for me.”* | Credible |
| Stigma and discrimination  *White et al., 2021* | *“They don’t give you a chance if you’re mentally ill.”* | Unequivocal |
| Discrimination  *Yang et al., 2023* | *“They called me a psychopath I did not have a name and people thought we were dangerous and would not associate with me let alone have sex with me.”* | Unequivocal |

*Fear of rejection*

| **Findings and citations** | **Illustrations** | **Level of credibility** |
| --- | --- | --- |
| The perceived risk of rejection is very high  *Budziszewska et al. 2020* | *“I’m afraid of rejection that if I were to get emotionally involved like it’s been many times before when I got involved with this or other guy and later got rejected. . .It was a very sad emotional state.”* | Unequivocal |
| Unrequited feelings  *Landi et al., 2020* | *“If you are also lovesick this is really a bad suffering… the gist of the issue is that at some point of time someone will fall in love with someone else if unrequited he will suffer.”* | Unequivocal |
| The risk of rejection  *White et al., 2021* | *“What I’m scared about most is rejection.”* | Unequivocal |

*Partner burden*

| **Findings and citations** | **Illustrations** | **Level of credibility** |
| --- | --- | --- |
| The emergence of psychosis can seriously affect mental well-being of the partner  *deJager et al., 2017* | *“He just really wanted to help me. Imagine seeing someone you love slip away and not being able to help. It was very hard for him. At a certain point he became very depressed. I felt and still feel extremely guilty about it.”* | Unequivocal |
| Relationship management difficulties  *Landi et al., 2020* | *“I lived with my partner for one year and everything was well… We had a good relationship but the first time I was admitted to hospital she came to visit me and said “I don’t love you anymore. I am leaving you.””* | Unequivocal |
| Pressures their partners faced to ensure their recovery  *Robertson et al., 2015* | *“There's a lot of pressure for him [ex-partner] probably not to a lot of pressure for him to not have any needs for himself … a lot of pressure for him not to be able to say “this is too much” … ’cause then he's not caring then he's not supportive.”* | Unequivocal |
| A sense of guilt over the impact of their mental health difficulties on partners  *Robertson et al., 2015* | *“The biggest barrier to recovery is actually getting over what you've done [to your partner] (nervous laughter) when you were unwell. Um … um … I think that was very hard.”* | Unequivocal |

*Sexual abuse experiences*

| **Findings and citations** | **Illustrations** | **Level of credibility** |
| --- | --- | --- |
| Risk of sexual abuse and need for stable relationships  *Budziszewska et al. 2020* | *“I mean he abused me sexually. I felt that he’s abusing me (. . .) Afterward I thought to myself if I were to find another man it would be a “white marriage” you know a relationship with no sex life. I was really determined to. . . I’ve been abstinent for. . . a year year and a half yeah.”* | Unequivocal |
| Sexual abuse  *deJager et al., 2017* | *“When I was younger I let people walk over me. Or I would keep pushing my own boundaries. Especially with boys I found it hard to say no. I kept wanting to please the other.”*  *More than one third of the sample (n=10, 36%) had experienced sexual abuse, particularly women (n=6, 67%; men n=4, 21%)* | Credible |
| Abusive relationships  *McCann 2010* | *“Sometimes Jack is a bit rough and I get sore. I get you know dry and he just forces it in. I tell him to stop but he just carries on.”* | Unequivocal |
| Desire to access talking therapy to process previous traumatic experiences  *Robertson et al., 2015* | *“I don’t talk to my workers about anything […] they don’t know anything about me past and they should do […] I don’t talk about anything about my past […] for what I’ve been through in my time it’s not an injection I need its counselling it’s more support more talkin’ about getting’ me past getting it out”*  *“Support with your relationships? Only so far as if they were able to identify where it wasn’t going well and was detrimental to your health to perhaps say: ‘Is this relationship working for you?’ to ask that kind of question.”* | Credible |

**Services are lacking and act as obstacles for service users to engage in romance and/or intimacy and its discussion**

*Romance and intimacy as too personal (lack of trust in staff), private, or taboo*

| **Findings and citations** | **Illustrations** | **Level of credibility** |
| --- | --- | --- |
| Romance as a private affair  *Forrester-Jones et al., 2023* | *“Care workers are people like anyone else [and] you’re kind of asking them for advice on romance. I don’t know. That’s their life as well. They’re gonna draw from their own life and knowledge and their romantic life might be private to them. I think it’s a bit tricky that kind of thing.”* | Unequivocal |
| Sexuality in the ward presented as crossing limits: reference to the topic  *Kaplan et al., 2022* | *“Really maybe if there was a little bit more reference to the topic [...] would be like healthy to this topic so maybe it would light things up more positively rather than being such a taboo.”* | Unequivocal |
| Shame of talking about sexual problems  *Raisi et al., 2018* | *"They may be ashamed in this regard and this is difficult for me to talk about."*  *"I can pull my carpet out of the water" [Iranian proverb: I do not need anyone else; I can do everything for myself].* | Credible |
| Power imbalance between themselves and mental health professionals: incongruent to discussing romantic relationships  *White et al., 2021* | *“I’d have to trust them […] that’s why I didn’t like about hospital cos there was so many people who were new and they didn’t know me an’ they were treatin’ me like a criminal […] it didn’t help me all they kept doin’ was punishing me givin’ more drugs.”* | Unequivocal |
| Privacy issues related to sex topics  *Yang et al., 2023* | *“I have never talked to others…I mean it is still too private.”* | Unequivocal |

*Lack of privacy or appropriate space*

| **Findings and citations** | **Illustrations** | **Level of credibility** |
| --- | --- | --- |
| Absence of a space in the facility to fulfil sexual needs  *Kaplan et al., 2022* | *“Even if I had asked there is no place. Will they provide us a private room? And will they allow us to get inside alone? They'll put cameras inside […] it's something basic that human beings need like a toilet that's the same thing that's hard.”*  *“I can fulfil my sexuality only if it feels intimate enough for me for example in the shower or under the blanket when everyone was sleep.”* | Credible |
| Discomfort from the small and uncomfortable environment  *Landi et al., 2020* | *“Here [in the facility] professionals have told me to use the bathroom for doing that [masturbation]. But it is not so comfortable especially having to stand up or seated on the lavatory.”* | Unequivocal |
| Concerns in institutional settings: no privacy  *McCann, 2010* | *“There is no privacy around here. There’s not much chance to have sex. We’re under the staff. Staff just come into the room they don’t bother to knock. I have no one to talk to about this stuff and I get worried that I may harm her. I feel anxious about it and it makes us both unhappy.”* | Unequivocal |
| A lack of privacy  *Quinn & Happell 2016* | *"I've seen people who have a wife or girlfriend and they visit and sit in the little waiting room and the girl has to give him a hand job or something… It would be nice if there was a room they could go to but there is nowhere else for them to go.”* | Unequivocal |
| Difficulties of finding hiding places for sex  *Quinn & Happell 2015a* | *“People now are having relationships and they have got to hide and that’s not very good. Perhaps there should be some rule where they could sleep together once a week or something like that in a flat or in a room or in their room. They shouldn’t have to hide.”* | Unequivocal |
| The absence of a private and dignified place  *Quinn & Happell 2015b* | *“I do know of people who find places on Campus to have sex but it's not a very comfortable situation it's not very intimate and you shouldn't have to go to those extremes of finding some secret spot it's not the best especially for people who might be a little paranoid a heightened sense it could be damaging to their mental state.”* | Unequivocal |
| A lack of private space  *Robertson et al., 2015* | *“Everyone else could come and just sit down beside you or the staff can just wander past and listen in … so there there is a real lack of private space … [later in transcript] there is never any real sense of you being able to have … the relationship.”* | Unequivocal |
| Lack of private spaces  *Yang et al., 2023* | *“Once I tried to masturbate in the bathroom but maybe I had been away from the room for too long so the nurse found me…So there there is a real lack of private space… I hope to be provided a warm and comfortable place.”* | Unequivocal |

*Lack of opportunity to form and maintain relationships: service environment*

| **Findings and citations** | **Illustrations** | **Level of credibility** |
| --- | --- | --- |
| Having sex under circumstances that are not only potentially risky but also likely be questioned as indicating a lack of sound judgement  *Brown et al., 2014* | *“Sex is an organised act that two people come together and do – and they’re going to do it wherever that is you know under a tree at the end of a tunnel they’re still going to do it… So I walked in and went to put my coat round there and they (two male patients) were having sex in the corner … and it’s not the first time they’d done that actually they’d done it somewhere else as well.”* | Unequivocal |
| Lack of places and spaces to meet a romantic partner  *Forrester-Jones et al., 2023* | *“Care in the community has huge cuts. Especially day centres. ‘Cause day centres - they give you a chance to mix and meet people and do group therapy like assertiveness training or anxiety management confidence building and [to] meet people. And that’s all gone.”* | Unequivocal |
| Regulation of sexual relations  *Landi et al., 2020* | *“Regarding love and affectivity one of the unwritten rules when you enter the facility is that having sexual intercourses is forbidden. It could happen that two lovers live together in the same facility however for the sake of other users it is not possible to have sex.”*  *“When X and Y kiss each other they annoy many. It is not tactful to kiss and touch genital parts in our presence.”* | Credible |
| Unusual environment that restricts many of the normal activities that a couple might engage in  *Quinn & Happell 2015a* | *“You’re very limited as to what you can do here. It’s not like you can go to the movies or go and see a band or anything like that. . . . Most people in a relationship get to have sex in the night time. This can never happen for us. We never as patients get to spend that time with each other so if you want to have sex it has to be during the day time and try and do it without getting caught.”* | Unequivocal |
| The absence of opportunity for romance  *Quinn & Happell 2015b* | *“In jails people get locked up for 20 years but they get conjugal visits with their partners some people get married in jail. Here there is nothing and it's wrong. The courts have the power to say we'll stay here for the rest of our lives. It's so lonely and miserable to have no one.”* | Unequivocal |
| Rapid looming environmental changes  *Yang et al., 2023* | *“Suddenly locked up in this closed environment I am not connected to anything personally or spiritually I am just kind of there.”* | Unequivocal |

*Lack of support from staff and services*

| **Findings and citations** | **Illustrations** | **Level of credibility** |
| --- | --- | --- |
| Exclusion of sexuality: forensic context  *Brown et al., 2014* | *“I find that they’re generally not approachable you know if it comes – you talk about the medication you talk about you know they’ve got a checklist of things that they want to talk about … And any deeper issues I find that I can’t talk to them about it. No. No … The deeper issues no …”* | Unequivocal |
| Lack of support from services for romantic relationships  *Forrester-Jones et al., 2023* | *“I found that there was very little [social care] support offered. But at the same time I wasn’t demanding much”.* | Credible |
| Nobody to talk to in the ward regarding sexual issues  *Kaplan et al., 2022* | *“It's because nobody cares. The doctor psychiatrist he asks other things entirely like 'Are you hearing voices?' […] There is only one doctor for twelve patients and he may talk to us every day but he doesn't really have the time to listen.”*  *“I’ve heard people talking about it with the staff with no problem.”* | Credible |
| Affectivity and sexuality was handled unsatisfactorily by practitioners  *Landi et al., 2020* | *“Sometimes [the mental health practitioners] give you the usual textbook answers in terms that are too specific to be understood properly. Moreover they judge [the expression of affection and sexuality] as frivolous and unimportant and this hurts a lot.”* | Unequivocal |
| Lack of support from care systems  *Ostman 2014* | *“Participants stressed that family members were generally on hand when needed for various everyday life situations as opposed to staff members who were usually harder to reach since they were engaged with other projects or patients.”*  *“Nowadays there is no sex in my life but many years ago when I was hospitalized I could meet partners at dances in the hospital.”* | Credible |
| Lack of support from nurses  *Quinn & Happell, 2016* | *“The staff tend to say no than yes even though you might be well and consent to it. We were consenting and really cared about each other we're adults yet we are told no and treated like children like it isn't normal or something.”* | Unequivocal |
| Negative experiences of services  *Robertson et al., 2015* | *“He pushed me against the wall and felt me up basically um…and I forget what he said something I kind of blanked it out a little bit I think he said something along the lines of “That's what you’re missing” or something and I actually reported that to the nurse … and nothing happened.”* | Unequivocal |
| The failure of services to ask about participants’ intimate relationships needs  *Robertson et al., 2015* | *“They are just looking at things from some psychiatrists not all from a biochemical viewpoint and that your brain is broken we’ll fix your brain with some medication and there you know there is no you know your social and personal life has very little to do with your your madness you know.”*  *“I had a male doctor and he when I brought that up the subject [sexual dysfunction] he just went red (laughs) really really red. Really red. He said “OK I’ll note it” and that was all was said about it.”* | Credible |
| The nature of the medical model approach underlying service delivery  *White et al., 2021* | *Appointments are typically brief intermittent and focused predominantly on symptoms rather than psychosocial issues - this was a barrier to effective therapeutic relationships.* | Unequivocal |

*Desire for advice and education on sexual intimacy, romantic relationships, and safe sex*

| **Findings and citations** | **Illustrations** | **Level of credibility** |
| --- | --- | --- |
| Measures for safe sex should be supplied to patients  *Kaplan et al., 2022* | *“One of the reasons that we don’t have sex is because there isn’t any protection.”* | Unequivocal |
| Educational programs and dedicated meetings concerning sexual and affective needs are lacking  *Landi et al., 2020* | *“I am self-taught: with my smartphone I read on internet about the treatments and the transmission of the [sexually transmitted] diseases that is how I keep myself updated.”*  *When asked about the possibility of receiving information on STD and unwanted pregnancy prevention reactions of RPFUs were discordant. Some argued that they did not see the need for it while others would be interested.* | Credible |
| Sexual knowledge and understanding lacking  *McCann 2010* | *“It started off with us being taught about the human body biology . . . male and female to say we received sexual education – no not really. Oh no nothing in the hospital it was never discussed.”* | Unequivocal |
| Sex education is not a priority  *Raisi et al., 2018* | *“I think that sexual issues are a part of normal human development and I do not feel the need to speak about this issue with physicians.”* | Unequivocal |
| Sexual education is often scarce or non-existent  *Yang et al., 2023* | *“To say we received sexual education—no not really. Oh no, no doctor or nurse has ever told us this; it was never discussed.”* | Unequivocal |

*Recommendations for communication within services on romance and intimacy*

| **Findings and citations** | **Illustrations** | **Level of credibility** |
| --- | --- | --- |
| Participants supported the notion that mental health teams could support their sexual health and wellbeing  *Brand et al., 2022* | *“I think the mental health team can improve my relationship but not sexual-wise yeah. You know more about communicating and all that probably.”*  *Five participants (of 14) either reported that mental health teams had no role to play or were uncertain how their mental health team could support them.* | Credible |
| Improvements in the hospital: creating groups  *Kaplan et al., 2022* | *“You need to create a group to talk about sexuality. And then to hear if a person speaks correctly to encourage them to keep going that way.”*  *“It’s intimate you don’t get into it in group sessions.”* | Credible |
| Mental health practitioners’ openness towards the theme  *Landi et al., 2020* | *“Yes I would like to talk about these things [sexual and affective needs] but I am shy and afraid of people. I know that if I could talk about it I may be helped.”*  *Most participants reported not having problems in disclosing their affective and sexual lives to MHPs* | Credible |
| Proposals for improving the way affective and sexual needs are addressed: professional-user talk time  *Landi et al., 2020* | *“I would like to propose a little change: that is to reserve professional–user times in which we can talk about affectivity sexuality and why not socialization in general which is an important thing itself.”* | Unequivocal |
| Expectations related to different kinds of support and sexual and relationship needs: responses were very varied  *McCann 2010* | *“I’m open to them asking me about it. If you asked me 20 years ago I’d probably be closeted.”*  *“They [health professionals] are only interested if you were considered abnormal. Like do you fancy kids or something like that then you would get a whole load of questions. I think they ask questions in a negative sense and that it was just one more mental box to cross off.”* | Unequivocal |
| Clinicians should be trained in communicating concerns  *Raisi et al., 2018* | *“My doctor told him (my fiancé) that he must be my caregiver for the whole of his life. He (The clinician) believed that I could not be a good mother. My fiancée wasn't worried before that. We had had a good relationship… after the session; he was afraid and really disappointed.”* | Unequivocal |
| Support groups: potentially did have utility  *White et al., 2021* | *“It could be a sort of group therapy session cos I really think that they can be very useful […] you’re meeting people that you can empathise with more potentially.”* | Unequivocal |
| Mental health professionals should adopt a sensitive approach when initiating conversations about relationships  *White et al., 2021* | *“Be sensitive to whether the person reacts by feeling to want to open up and discuss or wants to clam up shut down and say nothing and respect that.”* | Unequivocal |
| The importance of the therapeutic alliance  *White et al., 2021* | *“If they [mental health professionals] could be more sort of aware that there is a romantic aspect to a patient as well […] you’re not just a patient you’re not just a service user you have needs you’re a human being human beings need other people.”* | Unequivocal |
| It would be useful to have emotional support from nurses along with information and supportive advice  *Quinn & Happell 2016* | *“No the staff aren't supportive at all. I've had relationships in the hospital before and the staff weren't supportive at all and generally that is the reason I ended it because I got sick of all the stuff from staff. … if you're having any sexual problems it'd be good to be able to talk about it with someone and share your problems. But that never happens.”* | Unequivocal |
