## Supplementary material for "Systematic review and meta-aggregate analysis of mental health service users’ perspectives on romantic and intimate relationships and their support needs": S6 File. GRADE-CERQual ratings.

GRADE-CERQual ratings

**Overall rating**

**Instructions:**

Look across the assessments you have made for each CERQual component. Note particularly any components for which you have serious concerns.

• Decide whether you will ‘rate down’ (i.e., lower the level of confidence in the review finding) at all for the concerns identified and, if so, whether you will rate down by one or two levels. When making this overall assessment, consider the following:

 - Typically, the overall assessment of confidence should be rated down by at least one level for each component for which you have identified serious concerns

 - Where concerns in relation to a component are minor or moderate, it may not be necessary to rate down. However, if there are a number of such concerns, it may be appropriate to rate down by one level to represent two or more of these concerns

• When making a judgement on whether to ‘rate down’, also consider the following:

 - To some extent, the importance of concerns regarding a CERQual component needs to be judged in relation to the review finding. For instance, where a finding represents ‘mid-level’ theory regarding a phenomenon, it may be important that this is backed by considerable data and that the fit between the data from the primary studies and the review finding is clear. Concerns regarding adequacy of data and coherence may therefore be particularly critical in making an overall CERQual assessment for this finding

 - The data contributing to a review finding may come from studies that are assessed to have different levels of concern in relation to a CERQual component. This variation can be captured in three ways: (1) make a judgment that captures the highest level of concern for the component; (2) make a judgment that captures the lowest level of concern for the component; or (3) make a judgment that captures the “middle ground” for the component.

*All of above guidance from Lewin et al., 2018.*

**Ratings:**

***Individuals with mental health conditions have a desire for romantic and intimate relationships:***

- High confidence: There were no or very minor concerns regarding methodological limitations and adequacy. There were minor concerns regarding coherence as the category of “The desire for and complexities of parenthood” is vaguely defined and may be ambiguous when reading the synthesised finding title (makes up 3/14 findings). There were minor concerns related to relevance as the studies’ data had had partial relevance to the review question as diagnoses and study settings while inclusive of the review question did not encompass its entirety – any mental health condition and any mental health service setting.

***Mental health conditions directly hinder romantic and/or intimate relationships at the individual-level:***

- High confidence: There were no or very minor concerns across methodological limitations, coherence, and adequacy. Minor concerns regarding relevance as the data contributing to the synthesised finding were partially relevant. The data pertained predominantly to populations with serious mental illness, while the review question aimed to address individuals with any mental health condition. In addition, the settings were rarely community or outpatient based (5/12 studies were).

***Psychosocial factors impact romantic and/or intimate relationships:***

- Moderate confidence: Minor concerns regarding relevance as the data contributing to the review finding were partially relevant. The data were predominantly from populations of serious mental illness while the review question addresses any mental health condition. Additionally, settings were rarely community or outpatient based (4/10 studies). There were minor concerns regarding adequacy as 12.5% of papers had a small number of participants (6 ppt). Lastly, there were minor concerns regarding coherence as the finding ‘sexual abuse experiences’ is vaguely included in the synthesised finding’s description, with ambiguity surrounding whether this is an interpersonal factor (or could be explained by a different interpretation, e.g., a past life experience that is hindering pursuing romance and intimacy). This makes up 4/24 findings of which 2 were rated as credible certainty.

***Services are lacking and act as obstacles for service users to engage in romance and/or intimacy and its discussion:***

- Moderate confidence: There were moderate concerns regarding relevance as the data contributing to the review findings were partially relevant and some were unclear. 5 studies had a phenomena of interest that varied from the review question, 21 findings had studies with partial relevance as diagnoses and study settings while inclusive of the review question did not encompass its entirety – any mental health condition and any mental health service setting.

**Process**

**Assessment levels:**

- No or very minor concerns
- Minor concerns
- Moderate concerns
- Serious concerns

**Methodological limitations**

The extent to which there are problems in the design or conduct of the primary studies that contributed evidence to a review finding. For this item, ratings differing from ‘None or very minor concerns’ were defined as follows. Serious concerns: 50% or more of the studies were of low quality (score less than 4)*. Moderate concerns: 50% or more of the contributing studies were moderate or low quality (50% with score of less than 7). Minor concerns: some concerns exist with individual studies but not meeting criteria for moderate or serious concern defined above.

- See Critical Appraisal Quality Rating of the study associated with each of the aggregated findings

1. Individuals with mental health conditions have a desire for romantic and intimate relationships

Papers: 1, 2, 7, 7, 8, 12, 13, 15, 16, 16, 17

1: 8/10 (high). 2: 9/10 (high). 7: 9/10 (high). 8: 7/10 (high). 12: 8/10 (high). 13: 8/10 (high). 15: 9/10 (high). 16: 9/10 (high). 17: 8/10 (high).

- No or very minor concerns: all studies were rated as high quality.

2. Mental health conditions directly hinder romantic and/or intimate relationships at the individual-level

Papers: 1, 3, 3, 4, 5, 6, 7, 8, 8, 9, 9, 10, 15, 16, 17, 17, 17

1: 8/10 (high). 3: 10/10 (high). 4: 7/10 (high). 5: 9/10 (high). 6: 9/10 (high). 7: 9/10 (high). 8: 7/10 (high). 9: 9/10 (high). 10: 10/10 (high).15: 9/10 (high).16: 9/10 (high). 17: 8/10 (high).

- No or very minor concerns: all studies were rated as high quality.

3. Psychosocial factors impact romantic and/or intimate relationships

Papers: 1, 3, 3, 3, 4, 4, 4, 4, 5, 5, 7, 7, 8, 8, 8, 9, 9, 15, 15, 15, 16, 16, 16, 17, 17

1: 8/10 (high). 3: 10/10 (high). 4: 7/10 (high). 5: 9/10 (high). 6: 9/10 (high). 7: 9/10 (high). 8: 7/10 (high). 9: 9/10 (high). 14: 8/10 (high). 15: 9/10 (high).16: 9/10 (high). 17: 8/10 (high).

- No or very minor concerns: all studies were rated as high quality.

4. Services are lacking and act as obstacles for service users to engage in romance and/or intimacy and its discussion

Papers: 1, 2, 2, 5, 5, 6, 6, 6, 6, 7, 7, 7, 7, 7, 8, 8, 8, 10, 11, 11, 11, 12, 12, 13, 13, 14, 14, 15, 15, 16, 16, 17, 17, 17

1: 8/10 (high). 2: 9/10 (high). 5: 9/10 (high). 6: 9/10 (high). 7: 9/10 (high). 8: 7/10 (high). 10: 10/10 (high). 11: 8/10 (high). 12: 8/10 (high). 13: 8/10 (high). 14: 8/10 (high). 15: 9/10 (high). 16: 9/10 (high). 17: 8/10 (high).

- No or very minor concerns: all studies were rated as high quality.

**Coherence**

In essence, how clear and cogent the fit is between the data from the primary studies and a review finding that synthesises that data (88).

- Assess based on **a**) whether some of the data from included studies contradict the review finding (elements of the data from included studies might not fit the description of the key patterns captured in the finding); **b**) it is not clear/ambiguous if some of the underlying data support the review finding (e.g., vaguely defined or described); **c**) there are plausible alternative descriptions, interpretations or explanations could be used to synthesise the underlying data (is there different ways of describing, interpreting or explaining the data related to your finding?).

1. Individuals with mental health conditions have a desire for romantic and intimate relationships

Findings: 14 (13U 1C)

Papers: 1, 2, 7, 7, 8, 12, 13, 15, 16, 16, 17

- B) parenthood is not clearly understood from this synthesised finding description, but only makes up 3/14 findings [minor -> moderate] going with minor as it is a minority of findings

2. Mental health conditions directly hinder romantic and/or intimate relationships at the individual-level

Findings: 18 (18U)

Papers: 1, 3, 3, 4, 5, 6, 7, 8, 8, 9, 9, 10, 15, 16, 17, 17, 17

- A) Data related to impact of medication for the most part corroborates the finding that mental health conditions directly hinder but one participant from study 8 found antidepressants increased sex drive [no or very minor concerns -> minor concerns] as it is 1/18, going with no or very minor.

3. Psychosocial factors impact romantic and/or intimate relationships

Findings: 24 (16U 8C)

Papers: 1, 3, 3, 3, 4, 4, 4, 4, 5, 5, 7, 7, 8, 8, 8, 9, 9, 15, 15, 15, 16, 16, 16, 17, 17

- B and C) sexual abuse experiences is vaguely included in the review finding’s description, with ambiguity surrounding whether this is an interpersonal factor (or could be explained by a different interpretation, e.g., a past life experience that is hindering pursuing romance and intimacy). However, this makes up 4/24 findings of which 2 were rated as credible certainty [minor concern]

4. Services are lacking and act as obstacles for service users to engage in romance and/or intimacy and its discussion

Findings: 39 (29U 10C)

Papers: 1, 2, 2, 5, 5, 6, 6, 6, 6, 7, 7, 7, 7, 7, 8, 8, 8, 10, 11, 11, 11, 12, 12, 13, 13, 14, 14, 15, 15, 16, 16, 17, 17, 17

- No or very minor concerns

**Adequacy of the data**

An overall determination of the degree of richness and the quantity of data supporting a review finding. When assessing data adequacy, our aim is not to judge whether data adequacy has been achieved, but to judge whether there are grounds for concern regarding data adequacy that are serious enough to lower our confidence in the review finding (89).

- Assess the extent to which the information that the individual study authors have provided is detailed enough to allow the review author to interpret the meaning and context of what is being researched.
- Assess whether there is a sufficient number of studies and participants that the data related to a synthesised finding is coming from. Refer to the characteristics of studies table for studies’ sample sizes.

1. Individuals with mental health conditions have a desire for romantic and intimate relationships

- 1/11 (9.09%) papers had a small number of participants (6 ppt) [no or very minor]

Findings: 14 (13U 1C)

Papers: 1, 2, 7, 7, 8, 12, 13, *15*, 16, 16, 17

1: ppt = 13, 2: ppt = 20, 7: ppt = 13, 8: ppt = 30, 12: ppt = 10, 13: ppt = 10, 15: ppt = 6, 16: ppt = 10, 17: ppt = 20.

2. Mental health conditions directly hinder romantic and/or intimate relationships at the individual-level

- 1/17 (5.88%) papers that contributed to the findings had a small number of participants (6 ppt) [no or very minor]

Findings: 18 (18U)

Papers: 1, 3, 3, 4, 5, 6, 7, 8, 8, 9, 9, 10, *15*, 16, 17, 17, 17

1: ppt = 13, 3: ppt = 10, 4: ppt = 28, 5: ppt = 10, 6: ppt = 13, 7: ppt = 13, 8: ppt = 30, 9: ppt = 20, 10: ppt = 20, 15: ppt = 6, 16: ppt = 10, 17: ppt = 20.

3. Psychosocial factors impact romantic and/or intimate relationships

- 3/24 (12.5%) papers had a small number of participants (6 ppt) [minor concern]

Findings: 24 (16U 8C)

Papers: 1, 3, 3, 3, 4, 4, 4, 4, 5, 5, 7, 7, 8, 8, 8, 9, 9, 15, 15, 15, 16, 16, 16, 17, 17

1: ppt = 13, 3: ppt = 10, 4: ppt = 28, 5: ppt = 10, 7: ppt = 13, 8: ppt = 30, 9: ppt = 20, 15: ppt = 6, 16: ppt = 10, 17: ppt = 20.

4. Services are lacking and act as obstacles for service users to engage in romance and/or intimacy and its discussion

- 4/34 (11.76%) papers had a small number of participants (4 and 6 ppt) [no or very minor]

Findings: 39 (29U 10C)

Papers: 1, 2, 2, 5, 5, 6, 6, 6, 6, 7, 7, 7, 7, 7, 8, 8, 8, 10, 11, 11, 11, 12, 12, 13, 13, *14, 14, 15, 15*, 16, 16, 17, 17, 17

1: ppt = 13, 2: ppt = 20, 5: ppt = 10, 6: ppt = 13, 7: ppt = 13, 8: ppt = 30, 10: ppt = 20, 11: ppt = 10, 12: ppt = 10, 13: ppt = 10, 14: ppt = 4, 15: ppt = 6, 16: ppt = 10, 17: ppt = 20.

**Relevance**

The extent that the body of data from the primary studies from which the findings were extracted is applicable to the context (population, phenomenon of interest, setting) specified in the review question (90).

- See study characteristics table for details of each study related to the findings. Assess information in the following columns for each finding: Setting, Phenomena of interest, and Diagnoses of patients.
- Assess a) Indirectness: if a review domain, e.g. setting, perspective or population has been substituted by another in the primary study. E.g. experience of coercion of service users who are in hospital voluntarily, not involuntarily); b) Partiality: studies identify only some of the relevant review domains (e.g. data on one setting only, when review interest is broader); c) Uncertainty: the extent to which the focus of the included studies reflects the phenomenon of interest is uncertain, because of deficiencies in the reported details of the population, intervention, or settings. E.g. a study reports on involuntary treatment experiences, but also treatment experiences in general, and assessors are unsure to what extent results relate to involuntary experiences.
- In making judgements of the overall relevance, bare in mind that you’re not seeking a perfect fit between the included studies and the context of the review question. However, confidence in a review finding may, be reduced where the relation between the contexts of the primary studies and that specified in the review question is not apparent.

1. Individuals with mental health conditions have a desire for romantic and intimate relationships- [minor]

Findings: 14 (13U 1C)

Papers: 1, 2, 7, 7, 8, 12, 13, 15, 16, 16, 17

Indirectness:

- No or very minor concerns (3)

Uncertainty:

- No or very minor (3)
- Minor (2)
- Moderate (4)

Partiality:

- Minor (16)
- Moderate (1)

1:

- Partiality: setting is forensic unit and treated under forensic order explores only some of the relevant domains of this review (e.g., this review also explores community-based care) [minor]
- Uncertainty: phenomena of interest centres on sexuality and sexual experiences, making it somewhat difficult to distinguish the extent of which their results relate to romantic and/or intimate relationship needs [moderate]
- Partiality: diagnoses are schizophrenia, bipolar, unspecified psychosis which this review aimed to analyse but the review interest is broader (having a mental health condition and having had/currently be mental health service user) [minor]

2:

- Partiality: setting is forensic unit [minor]
- Uncertainty: phenomena of interest centres on exclusion of sexuality [moderate]
- Partiality: diagnoses of a psychotic disorder, severe depression [minor]

7:

o	Partiality: setting [minor]

o	Uncertainty: phenomena of interest is to better understand how sexuality and affectivity are expressed by service users within a residential setting [moderate]

o	Partiality: diagnoses [minor]

8:

- Partiality: setting was a clinic in London that provided antipsychotic depot injections [moderate]
- Uncertainty: phenomena of interest is to capture in-depth perspectives of people with a medical diagnosis of schizophrenia regarding intimate relationships [none or very minor]
- Partiality: Schizophrenia, schizotypal and delusional disorder [minor]

12:

- Partiality: setting was a secure forensic hospital [minor]
- Uncertainty: phenomena of interest was to elicit the views of consumers and nurses about forming sexual relationships within this long-term and secure setting [none or very minor]
- Indirectness: does not specify diagnoses [no or very minor]

13:

- Partiality: setting was a secure forensic hospital [minor]
- Uncertainty: phenomena of interest to explore perceptions of privacy and dignity for sexual relationships in a forensic mental health hospital t forming sexual relationships within this long-term and secure setting [minor]
- Indirectness: does not specify diagnoses [none or very minor]

15:

- Partiality: setting was an inpatient psychiatric hospital [minor]
- Partiality: phenomena of interest was to explore the psychiatric inpatient experiences of lesbian and gay service users in relation to their intimate relationship needs and how these experiences affected their mental health recovery [minor]
- Partiality: diagnoses [minor]

16:

- Partiality: setting was a community-based mental healthcare services [minor]
- Partiality: phenomena of interest was to investigate how people with experience of psychosis conceptualise romantic relationships and what support they would like in this area of their lives [none or very minor]
- Partiality: diagnoses – experience of psychoses [minor]

17:

- Partiality: setting was inpatient and outpatient psychiatric hospital [minor]
- Partiality: phenomena of interest was to explore the sexual needs of people with schizophrenia and identify factors hindering sexual activities [none or very minor]
- Partiality: diagnoses – schizophrenia [minor]

2. Mental health conditions directly hinder romantic and/or intimate relationships at the individual-level [minor]

Findings: 18 (18U)

Papers: 1, 3, 3, 4, 5, 6, 7, 8, 8, 9, 9, 10, 15, 16, 17, 17, 17

Indirectness:

- No or very minor concerns: 2

Partiality:

- No or very minor concerns: 3
- Minor concerns: 19
- Moderate concerns: 2

Uncertainty:

- No or very minor concerns: 5
- Minor concerns: 2
- Moderate concerns: 2
- Serious concerns: 1

1:

- Partiality: forensic unit and treated under forensic order explores only some of the relevant domains of this review (e.g., this review also explores community-based care) [minor]
- Uncertainty: phenomena of interest centres on sexuality and sexual experiences, making it somewhat difficult to distinguish the extent of which their results relate to romantic and/or intimate relationship needs [moderate]
- Partiality: diagnoses are schizophrenia, bipolar, unspecified psychosis which this review aimed to analyse but the review interest is broader (having a mental health condition and having had/currently be mental health service user) [minor]

3:

- Partiality: setting is inpatient and outpatient psychiatric clinic [no or very minor concerns]
- Uncertainty: phenomena of interest examines how the illness has influenced their emotional experiences regarding love and their intimate relationship experiences, with an explicit intention to give voice to the patient’s perspective [minor]
- Partiality: diagnoses of schizophrenia [minor]

4:

- Partiality: setting is community mental health care [minor]
- Uncertainty: phenomena of interest examines which problems participants encounter in establishing intimacy and maintaining intimate relationships [no or very minor]
- Indirectness: does not specify diagnoses [no or very minor]

5:

- Partiality: setting is mental health charities and a mental health self-help network [minor]
- Uncertainty: phenomena of interest explores how people with mental health problems experience romantic relationships and what support they might need [no or very minor]
- Indirectness: does not specify diagnoses [no or very minor]

6:

- Partiality: setting is a psychiatric unit [minor]
- Uncertainty: phenomena of interest is to examine the sexual needs and challenges of people with severe mental illnesses admitted to an isolated psychiatric ward [no or very minor]
- Partiality: diagnoses were schizophrenia, psychotic disorders, or bipolar disorder [minor]

7:

- Partiality: setting was psychiatric residential setting [minor]
- Uncertainty: phenomena of interest is to better understand how sexuality and affectivity are expressed by service users within a residential setting [moderate]
- Partiality: diagnoses were psychotic disorders, personality disorders, bipolar disorder, depression [minor]

8:

- Partiality: setting was a clinic in London that provided antipsychotic depot injections [moderate]
- Uncertainty: phenomena of interest is to capture in-depth perspectives of people with a medical diagnosis of schizophrenia regarding intimate relationships [none or very minor]
- Partiality: Schizophrenia, schizotypal and delusional disorder [minor]

9:

- Partiality: setting was a psychosocial recovery centre [minor]
- Uncertainty: phenomena of interest was to better understand women with serious mental illness's experiences with romantic and intimate relationships [moderate]
- Partiality: Major depressive disorder, PTSD, anxiety, bipolar, borderline personality disorder, schizophrenia, schizoaffective [none or very minor]

10:

- Partiality: setting was an outpatient psychiatric services [minor]
- Uncertainty: phenomena of interest was to determine how people with serious mental illness experience sex and assess satisfaction with it in a broader evaluation of quality of life [serious]
- Partiality: diagnoses - all were currently in treatment in psychiatric out-patient services with serious mental illness [none or very minor]

15:

- Partiality: setting was an inpatient psychiatric hospital [minor]
- Partiality: phenomena of interest was to explore the psychiatric inpatient experiences of lesbian and gay service users in relation to their intimate relationship needs and how these experiences affected their mental health recovery [minor]
- Partiality: diagnoses [minor]

16:

- Partiality: setting was a community-based mental healthcare services [minor]
- Partiality: phenomena of interest was to investigate how people with experience of psychosis conceptualise romantic relationships and what support they would like in this area of their lives [none or very minor]
- Partiality: diagnoses – experience of psychoses [minor]

17:

- Partiality: setting was inpatient and outpatient psychiatric hospital [minor]
- Partiality: phenomena of interest was to explore the sexual needs of people with schizophrenia and identify factors hindering sexual activities [none or very minor]
- Partiality: diagnoses – schizophrenia [minor]

3. Psychosocial factors impact romantic and/or intimate relationships – [minor]

Findings: 24 (16U 8C)

Papers: 1, 3, 3, 3, 4, 4, 4, 4, 5, 5, 7, 7, 8, 8, 8, 9, 9, 15, 15, 15, 16, 16, 16, 17, 17

Indirectness:

- No or very minor: 2

Uncertainty:

- No or Very Minor Concerns: 6
- Minor Concerns: 3
- Moderate Concerns: 5

Partiality:

- No or Very Minor Concerns: 1
- Minor Concerns: 17
- Moderate Concerns: 2

1:

- Partiality: forensic unit and treated under forensic order explores only some of the relevant domains of this review (e.g., this review also explores community-based care) [minor]
- Uncertainty: phenomena of interest centres on sexuality and sexual experiences, making it somewhat difficult to distinguish the extent of which their results relate to romantic and/or intimate relationship needs [moderate]
- Partiality: diagnoses are schizophrenia, bipolar, unspecified psychosis which this review aimed to analyse but the review interest is broader (having a mental health condition and having had/currently be mental health service user) [minor]

3:

- Partiality: setting is inpatient and outpatient psychiatric clinic [no or very minor concerns]
- Uncertainty: phenomena of interest examines how the illness has influenced their emotional experiences regarding love and their intimate relationship experiences, with an explicit intention to give voice to the patient’s perspective [minor]
- Partiality: diagnoses of schizophrenia [minor]

4:

- Partiality: setting is community mental health care [minor]
- Uncertainty: phenomena of interest examines which problems participants encounter in establishing intimacy and maintaining intimate relationships [no or very minor]

5:

- Indirectness: does not specify diagnoses [no or very minor]
- Partiality: setting is mental health charities and a mental health self-help network [minor]
- Uncertainty: phenomena of interest explores how people with mental health problems experience romantic relationships and what support they might need [no or very minor]
- Indirectness: does not specify diagnoses [no or very minor]

6:

- Partiality: setting is a psychiatric unit [minor]
- Uncertainty: phenomena of interest is to examine the sexual needs and challenges of people with severe mental illnesses admitted to an isolated psychiatric ward [no or very minor]
- Partiality: diagnoses were schizophrenia, psychotic disorders, or bipolar disorder [minor]

7:

- Partiality: setting was psychiatric residential setting [minor]
- Uncertainty: phenomena of interest is to better understand how sexuality and affectivity are expressed by service users within a residential setting [moderate]
- Partiality: diagnoses [minor]

8:

- Partiality: setting was a clinic in London that provided antipsychotic depot injections [moderate]
- Uncertainty: phenomena of interest is to capture in-depth perspectives of people with a medical diagnosis of schizophrenia regarding intimate relationships [none or very minor]
- Partiality: Schizophrenia, schizotypal and delusional disorder [minor]

9:

- Partiality: setting was a psychosocial recovery centre [minor]
- Uncertainty: phenomena of interest was to better understand women with serious mental illness's experiences with romantic and intimate relationships [moderate]
- Partiality: Major depressive disorder, PTSD, anxiety, bipolar, borderline personality disorder, schizophrenia, schizoaffective [none or very minor]

14:

- Partiality: setting was an inpatient psychiatric hospital [minor]
- Uncertainty: phenomena of interest was to explore views of patients with severe mental illness and their care providers about sharing sexual problems with care providers in these patients within the context of Iran [moderate]
- Partiality: serious mental illness [none or very minor]

15:

- Partiality: setting was an inpatient psychiatric hospital [minor]
- Partiality: phenomena of interest was to explore the psychiatric inpatient experiences of lesbian and gay service users in relation to their intimate relationship needs and how these experiences affected their mental health recovery [minor]
- Partiality: diagnoses [minor]

16:

- Partiality: setting was a community-based mental healthcare services [minor]
- Partiality: phenomena of interest was to investigate how people with experience of psychosis conceptualise romantic relationships and what support they would like in this area of their lives [none or very minor]
- Partiality: diagnoses – experience of psychoses [minor]

17:

- Partiality: setting was inpatient and outpatient psychiatric hospital [minor]
- Partiality: phenomena of interest was to explore the sexual needs of people with schizophrenia and identify factors hindering sexual activities [none or very minor]
- Partiality: diagnoses – schizophrenia [minor]

4. Services are lacking and act as obstacles for service users to engage in romance and/or intimacy and its discussion– [moderate]

Moderate concerns (5 studies with uncertain relevance as phenomena of interest varied from the review question, 21 studies had partial relevance as diagnoses and study settings were inclusive of the review question but not wholly)

Findings: 39 (29U 10C)

Papers: 1, 2, 2, 5, 5, 6, 6, 6, 6, 7, 7, 7, 7, 7, 8, 8, 8, 10, 11, 11, 11, 12, 12, 13, 13, 14, 14, 15, 15, 16, 16, 17, 17, 17

Indirectness:

- No or very minor (5)
- Minor (1)

Uncertainty:

- No or very minor (6)
- Minor (1)
- Moderate (4)
- Serious (1)

Partiality:

- No or very minor concerns (2)
- Minor (20)
- Moderate (1)

1:

- Partiality: forensic unit and treated under forensic order explores only some of the relevant domains of this review (e.g., this review also explores community-based care) [minor]
- Uncertainty: phenomena of interest centres on sexuality and sexual experiences, making it somewhat difficult to distinguish the extent of which their results relate to romantic and/or intimate relationship needs [moderate]
- Partiality: diagnoses are schizophrenia, bipolar, unspecified psychosis which this review aimed to analyse but the review interest is broader (having a mental health condition and having had/currently be mental health service user) [minor]

2:

- Partiality: setting is forensic unit [minor]
- Uncertainty: phenomena of interest centres on exclusion of sexuality [moderate]
- Partiality: diagnoses of a psychotic disorder, severe depression [minor]

5:

- Partiality: setting is mental health charities and a mental health self-help network [minor]
- Uncertainty: phenomena of interest explores how people with mental health problems experience romantic relationships and what support they might need [no or very minor]
- Indirectness: does not specify diagnoses [no or very minor]

6:

- Partiality: setting is a psychiatric unit [minor]
- Uncertainty: phenomena of interest is to examine the sexual needs and challenges of people with severe mental illnesses admitted to an isolated psychiatric ward [no or very minor]
- Partiality: psychotic disorders, personality disorders, bipolar, depression [minor]

7:

o	Partiality: setting [minor]

o	Uncertainty: phenomena of interest is to better understand how sexuality and affectivity are expressed by service users within a residential setting [moderate]

o	Partiality: diagnoses [minor]

8:

- Partiality: setting was a clinic in London that provided antipsychotic depot injections [moderate]
- Uncertainty: phenomena of interest is to capture in-depth perspectives of people with a medical diagnosis of schizophrenia regarding intimate relationships [none or very minor]
- Partiality: Schizophrenia, schizotypal and delusional disorder [minor]

10:

- Partiality: setting was an outpatient psychiatric services [minor]
- Uncertainty: phenomena of interest was to determine how people with serious mental illness experience sex and assess satisfaction with it in a broader evaluation of quality of life [serious]
- Partiality: diagnoses - all were currently in treatment in psychiatric out-patient services with serious mental illness [none or very minor]

11:

- Partiality: setting was secure forensic hospital [minor]
- Indirectness: phenomena of interest was to explore perceptions of nurses and patients regarding sexual intimacy in a long-term mental health unit [minor]
- Uncertainty: does not specify diagnoses [none or very minor]

12:

- Partiality: setting was a secure forensic hospital [minor]
- Uncertainty: phenomena of interest was to elicit the views of consumers and nurses about forming sexual relationships within this long-term and secure setting [none or very minor]
- Indirectness: does not specify diagnoses [none or very minor]

13:

- Partiality: setting was a secure forensic hospital [minor]
- Uncertainty: phenomena of interest to explore perceptions of privacy and dignity for sexual relationships in a forensic mental health hospital t forming sexual relationships within this long-term and secure setting [minor]
- Indirectness: does not specify diagnoses [none or very minor]

14:

- Partiality: setting was an inpatient psychiatric hospital [minor]
- Uncertainty: phenomena of interest was to explore views of patients with severe mental illness and their care providers about sharing sexual problems with care providers in these patients within the context of Iran [moderate]
- Partiality: serious mental illness [none or very minor]

15:

- Partiality: setting was an inpatient psychiatric hospital [minor]
- Partiality: phenomena of interest was to explore the psychiatric inpatient experiences of lesbian and gay service users in relation to their intimate relationship needs and how these experiences affected their mental health recovery [minor]
- Partiality: diagnoses [minor]

16:

- Partiality: setting was a community-based mental healthcare services [minor]
- Partiality: phenomena of interest was to investigate how people with experience of psychosis conceptualise romantic relationships and what support they would like in this area of their lives [none or very minor]
- Partiality: diagnoses – experience of psychoses [minor]

17:

- Partiality: setting was inpatient and outpatient psychiatric hospital [minor]
- Partiality: phenomena of interest was to explore the sexual needs of people with schizophrenia and identify factors hindering sexual activities [none or very minor]
- Partiality: diagnoses – schizophrenia [minor]
