## Supplementary material for "Systematic review and meta-aggregate analysis of mental health service users’ perspectives on romantic and intimate relationships and their support needs": S7 File. PROSPERO Protocol.

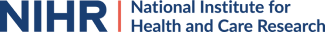

**Systematic review of mental health service users'**

**perspectives on romantic and intimate relationships**

**and their support needs.**

*Aisling Smith O’Connor,* *Helen Killaspy, Sharon Eager, Eunice Wu, Brynmor Lloyd-Evans.*

**Citation**

Aisling Smith O Connor, *Helen Killaspy, Sharon Eager, Eunice Wu, Brynmor Lloyd-Evans*. Systematic review of mental health service users' perspectives on romantic and intimate relationships and their support needs.

PROSPERO 2024 Available from

[htt](https://www.crd.york.ac.uk/PROSPERO/view/CRD42024503997)

[ps://www.crd.](https://www.crd.york.ac.uk/PROSPERO/view/CRD42024503997)

[y](https://www.crd.york.ac.uk/PROSPERO/view/CRD42024503997)

[ork.ac.uk/PROSPERO/view/CRD4202450399](https://www.crd.york.ac.uk/PROSPERO/view/CRD42024503997)

[7](https://www.crd.york.ac.uk/PROSPERO/view/CRD42024503997)

**PROSPERO**

International prospective register of systematic reviews

### REVIEW TITLE AND BASIC DETAILS

#### Review title

Systematic review of mental health service users' perspectives on romantic and intimate relationships and their support needs.

#### Review objectives

What are the views of mental health service users on their needs for romantic and intimate relationships, and how they can be best supported by mental health staff.

### SEARCHING AND SCREENING

#### Searches

The following electronic databases will be searched: MEDLINE (ovid), PsycINFO (ovid), CINAHL, and Web of Science.

Grey literature will also be explored using google search of “mental health” and

“romantic/intimate relationships” with a minimum of the first 10 result pages being explored, carrying on until no relevant papers are being retrieved.

Search strategy: The search strategy was designed to underpin the fundamental components of the research objective, yielding three key concepts: romantic and intimate relationships; mental health; and qualitative research. Synonyms for these core elements will be generated and combined using Boolean operators to create search. The searches will be set to identify studies that include all key concepts in the title or abstract of the study. Necessary adjustments to the search will be made to adapt to the specifications of each database. There will be no

restrictions on date of publication. However, the search will be limited to English peer-reviewed publications and fully available published articles. The searches will be re-run just before the final analyses with further studies being retrieved and included, if applicable.

Search concepts:

1) Romantic and intimate relationships

AND

2) Mental health

AND

3) Qualitative

#### Study design

This systematic review will encompass qualitative studies, incorporating a range of methodologies such as interviews, focus groups, and open-ended surveys.

### ELIGIBILITY CRITERIA

#### Condition or domain being studied

Intimate relationships defined by sexual activity, romantic love, or increased physical intimacy, encompassing monogamous and non-monogamous relationships.

#### Population

People with a mental health condition who have received/are receiving care and support from mental health services.

#### Intervention(s) or exposure(s)

Eligible papers will explore mental health service users’ needs for romantic/intimate relationships and how they feel mental health staff can best discuss this and offer support.

**Comparator(s) or control(s)** Not applicable

#### Context

The qualitative data can be generated from individuals with current or past experience of mental health services provided by the NHS, Local Authority, independent/private or voluntary sector.

### OUTCOMES TO BE ANALYSED

#### Main outcomes

To explore mental health service users’ views on their needs for and perspectives on receiving support from services with romantic/intimate relationships.

**Additional outcomes**

Not applicable

### DATA COLLECTION PROCESS

#### Data extraction (selection and coding)

Study selection: Covidence will be used to import the search results and will manage, compile, and evaluate the gathered studies. Reviewers will screen titles and abstracts for inclusion and reasons for exclusion will be noted – 10% of these screenings will undergo a double-screening process, conducted independently by two blinded raters. On completion of the screening stage, the remaining full texts will be reviewed by two independent raters. It may be necessary to communicate with study investigators to clarify research eligibility. Any disagreements between the two raters will be resolved through discussion or by consulting a third reviewer. The selection process will be documented with sufficient details necessary to create a PRISMA flowchart being recorded.

Data extraction: a form will be created and used to synthesize information from the selected studies. This will include first author, publication year, country, sample characteristics (e.g. age range, mean age, mental illness diagnosis), study setting, methods of data collection used (e.g.

focus groups), methods of data analysis (grounded theory, phenomenology, thematic, etc.), summary of the study’s aim/findings (key themes or concepts identified in the study), and quality of the study.

#### Risk of bias (quality) assessment

The JBI Critical Appraisal Checklist for Qualitative Research will be utilised to assess and note the methodological quality of included studies. It has been selected as it is a standardized critical appraisal instrument from the Joanna Briggs Institute System for the Unified Management, Assessment and Review of Information. Studies scoring seven out of ten and above will be classed as high quality, five to seven as medium quality, and those with a score of less than five as low quality.

An appraisal of the level of confidence in the findings from the review will also be conducted.

The overall assessment of the qualitative evidence synthesis will be assessed by the GRADECERQual with confidence in each of our synthesised findings being rated as high, moderate, low, and very low. This rating is made on the basis of methodological limitations, coherence, adequacy, and relevance.

### PLANNED DATA SYNTHESIS

#### Strategy for data synthesis

A meta-aggregative approach to the synthesis of qualitative evidence will be employed. This synthesis approach prioritises the practicality and usability of the primary author's findings, limiting subjectivity and preserving the original interpretations of the qualitative data, seeking to enable generalisable statements in the form of recommendations to guide practitioners and policy makers.

1. Extraction of all findings from all included studies with an accompanying illustration (e.g.

quotation) and allocated level of plausibility for each finding.

This will involve repeated reading of the text to identify relevant findings.

2. Developing categories for findings with at least two findings per category.

This will involve grouping two or more assembled findings based on similarity in meaning, following repeated, detailed examination of the extracted findings. The categories will include a brief description of the key concept that conveys the wholistic meaning of the group of similar findings.

3. Developing one or more synthesized findings of at least two categories.

This will be expressed as ‘indicatory’ statements, comprising an over-arching description of two or more categories.

#### Analysis of subgroups or subsets

No formal subgroup or comparative analyses are planned. However, during data synthesis we will consider from which clinical groups and service settings the findings are based, and any apparent differences in perspectives between groups will be documented.

### REVIEW AFFILIATION, FUNDING AND PEER REVIEW

#### Review team members

- Aisling Smith O’Connor, UCL
- Helen Killaspy, UCL
- Sharon Eager, UCL
- Eunice Wu, UCL
- Brynmor Lloyd-Evans, UCL

**Review affiliation**

UCL

**Funding source**

Not applicable

#### Named contact

Aisling Smith O Connor. University College London, Gower Street.

Brynmor Lloyd-Evans, University College London, Gower Street,

### TIMELINE OF THE REVIEW

**Review timeline**

Start date: 24 March 2024. End date: 30 August 2024

**Date of first submission to PROSPERO** 13 March 2024

**Date of registration in PROSPERO** 14 March 2024

### CURRENT REVIEW STAGE

**Publication of review results**

The intention is to publish the review once completed.The review will be published in English

#### Stage of the review at this submission

**Review stage	Started	Completed**

Pilot work

Formal searching/study identification

Screening search results against inclusion criteria

**Review stage	Started	Completed**

Data extraction or receipt of IP

Risk of bias/quality assessment

Data synthesis

**Review status**

The review is currently planned or ongoing.

### ADDITIONAL INFORMATION

#### PROSPERO version history

Version 1.0 published on 14 Mar 2024

#### Review conflict of interest None known

**Country** England

**Medical Subject Headings**

Humans; Mental Health; Mental Health Services; Sexual Behavior; Sexual Partners

#### Disclaimer

The content of this record displays the information provided by the review team. PROSPERO does not peer review registration records or endorse their content.

PROSPERO accepts and posts the information provided in good faith; responsibility for record content rests with the review team. The owner of this record has affirmed that the information provided is truthful and that they understand that deliberate provision of inaccurate information may be construed as scientific misconduct.

PROSPERO does not accept any liability for the content provided in this record or for its use.

Readers use the information provided in this record at their own risk.

Any enquiries about the record should be referred to the named review contact
